# An explainable AI framework for interpretable biological age

**DOI:** 10.1101/2022.10.05.22280735

**Authors:** Wei Qiu, Hugh Chen, Matt Kaeberlein, Su-In Lee

## Abstract

**Background:** An individual’s biological age is a measurement of health status and provides a mechanistic understanding of aging. *Age clocks* estimate a biological age of an individual based on their various *features*. Existing clocks have key limitations caused by the undesirable tradeoff between accuracy (i.e., predictive performance for chronological age or mortality, often achieved by complex, black-box models) and interpretability (i.e., the contributions of features to biological age). Here, we present ‘ENABL (<u>E</u>xplai<u>NA</u>ble <u>B</u>io<u>L</u>ogical) Age’, a computational framework that combines machine learning (ML) models with explainable AI (XAI) methods to accurately estimate biological age with individualized explanations.

**Methods:** To construct ENABL Age clock, we first need to predict an age-related outcome of interest (e.g., all-cause or cause-specific mortality), and then rescale the predictions nonlinearly to estimate biological age. We trained and evaluated the ENABL Age clock using the UK Biobank (501,366 samples with 825 features) and NHANES 1999-2014 (47,084 samples with 158 features) datasets. To explain the ENABL Age clock, we extended existing XAI methods so we could linearly decompose any individual’s ENABL Age into contributing risk factors. To make ENABL Age clock broadly accessible, we developed two versions: (1) ENABL Age-L, which is based on popular blood tests, and (2) ENABL Age-Q, which is based on questionnaire features. Finally, when we created ENABL Age clocks based on predictions of different age-related outcomes, we validated that each one captures sensible, yet disparate aging mechanisms by performing GWAS association analyses.

**Findings:** Our results indicate that ENABL Age clocks successfully separate healthy from unhealthy aging individuals and are stronger predictors of mortality than existing age clocks. We externally validated our results by training ENABL Age clocks on UK Biobank data and testing on NHANES data. The individualized explanations that reveal the contribution of specific features to ENABL Age provide insights into the important features for biological age. Association analysis with risk factors and aging-related morbidities, and genome-wide association study (GWAS) results on ENABL Age clocks trained on different mortality causes show that each one captures sensible aging mechanisms.

**Interpretation:** We developed and validated a new ML and XAI-based approach to calculate and interpret biological age based on multiple aging mechanisms. Our results show strong mortality prediction power, interpretability, and flexibility. ENABL Age takes a consequential step towards accurate interpretable biological age prediction built with complex, high-performance ML models.

**Research in context:** 

**Evidence before this study:** Biological age plays an important role to understanding the mechanisms underlying aging. We search PubMed for original articles published in all languages with the terms “biological age” published until June 22, 2022. Most prior studies focus on the first generation of biological age clocks that are designed to predict chronological age. These clocks have weak and variable associations with mortality risk and other aging outcomes. Only a few studies present the second-generation of biological age clocks, which are built directly with aging outcomes. However, these studies use linear models and do not provide individualized explanations. Moreover, previous biological age clocks cannot specify what aging process they capture. Unlike our study, none of the previous studies have combined a complex machine learning (ML) model and an explainable artificial intelligence (XAI) method, which allows us to build biological ages that are both accurate and interpretable.

**Added value of this study:** In this study, we present ENABL Age, a new approach to estimate and understand biological age that combines complex ML models and XAI method. The ENABL Age approach is designed to measure second-generation biological age clocks by directly predicting age-related outcomes. Our results indicate that ENABL Age accurately reflects individual health status. We also introduce two variants of ENABL Age clocks: (1) ENABL Age-L, which takes popular blood tests as inputs (usable by medical professionals), and (2) ENABL Age-Q, which takes questionnaire features as inputs (usable by non-professional healthcare consumers). We extend existing XAI methods to calculate the contributions of input features to ENABL Age estimate in units of years, which makes our biological age clocks more human-interpretable. Our association analysis and GWAS results show that ENABL Age clocks trained on different age-related outcomes can capture different aging mechanisms.

**Implications of all the available evidence:** We develop and validate a new ML and XAI-based approach to measure and interpret biological age based on multiple aging mechanisms. Our results demonstrate that ENABL age has strong mortality prediction power, is interpretable, and is flexible. ENABL Age takes a consequential step towards applying XAI to interpret biological age models. Its flexibility allows for many future extensions to omics data, even multi-omic data, and multi-task learning.

## 1 Introduction

Aging contributes to many diseases that affect all organ systems and is the leading factor associated with physical, cognitive and degenerative disorders, such as heart disease, neurodegeneration and cancer [38]. Measuring the state of aging (i.e., biological age) is the first step towards addressing age-associated disease and so extending lifespans.

*Biological age* measures the level of biological functioning of an organism, organ or cell as assessed in comparison to an expected level of function for a given chronological age [42] and reflects an individual’s general health status. The first generation of biological age clocks were designed to predict chronological age [16, 18, 49, 11] but have weak and variable associations with mortality risk and other aging outcomes [5, 19, 12]. Furthermore, extremely accurate prediction of chronological age can actually reduce the association of biological age with mortality risk [53], suggesting that chronological age predictors may overfit to “noise” unrelated to aging phenotypes [28, 42]. These findings led to the second-generation of biological age clocks, which are built directly with aging outcomes such as mortality [25, 26, 31, 51]. By design, second-generation biological ages have robust associations with mortality risk and other aging traits (e.g., heart disease risk, physical functioning and blood chemistry markers).

Existing biological age clocks have three main limitations. First, they necessitate a tradeoff between accuracy (i.e., predictive performance for chronological age or mortality, often achieved by complex, blackbox models) and interpretability (i.e., the contribution of each feature to the prediction). Most of them are linear models, which are easy to interpret but have weak predictive power. This choice is natural given that interpretability is a key goal of biological age clocks: identifying biomarkers of biological age can improve our understanding of the aging process and help develop drugs that target aging-related dysfunction. In contrast, recent work uses complex machine learning (ML) models to build biological age clocks. These clocks, while more accurate at predicting chronological age, are hard to interpret [17, 14, 43]. *To build models that are both accurate and interpretable, we turn to the emerging area of explainable AI* (XAI) [41, 32, 46].

The second limitation is that interpretations of previous biological age clocks may fail to address important scientific questions. In particular, linear models are typically interpreted based on the coefficients of features, which only explain the model as a whole (global explanation). However, the aging process varies widely for each individual, so *we leverage recent XAI methods to provide principled individualized (local) explanations based on feature attributions*. Typically, feature attributions can also be difficult for non-ML practitioners to understand because they provide explanations in units of predicted probability, or logits. To further extend the accessibility of ENABL Age explanations, *we rescale our attributions to the age scale in units of years* so that the rescaled attributions sum to an individual’s biological age acceleration.

The third limitation of current biological age clocks is that they cannot specify which aging process they are capturing. This is problematic because biological aging is enormously complex and thought to be driven by many biological processes [30, 22]. Previous studies found low agreement between biological age clocks in terms of both their correlations with each other and their relative associations with aging traits [1, 27, 8], suggesting that they may be measuring different aspects of biological age. To address this, it is important to distinguish among the multiple aging mechanisms that biological age captures. For this purpose, *we built our biological age clocks by predicting different age-related outcomes so we can specify the specific underlying aging mechanisms our clocks capture*.

To overcome the aforementioned limitations of prior biological age clocks, we present ENABL (ExplaiNAble BioLogical) Age, a new approach to estimating and interpreting biological age that combines complex ML and XAI methods. Our approach is designed to measure the second-generation biological age: it directly predicts an age-related outcome (e.g., all-cause mortality or cause-specific mortality) of interest and then nonlinearly rescales the predictions into units of age. We apply ENABL Age approach to UK Biobank (UKB; 501,366 samples with 825 features) and NHANES 1999-2014 (47,084 samples with 158 features) datasets. Our results indicate that ENABL Age successfully separates healthy from unhealthy aging individuals and is a stronger predictor of mortality than existing age clocks, which demonstrate that ENABL Age accurately reflects individual health status (Section 3.2). To make ENABL Age clock broadly accessible, we also introduce two variants of ENABL Age clocks: (1) ENABL Age-L, which takes popular blood tests as inputs (usable by medical professionals), and (2) ENABL Age-Q, which takes questionnaire features as inputs (usable by non-professional healthcare consumers). We ensure generalizability by externally validating ENABL Age clock trained on UKB data using the NHANES data. We extend existing XAI methods to calculate the contributions of ENABL Age input features in units of years, which makes our biological age clocks more human-interpretable than traditional explanations, which are presented in terms of predicted probability, or logits (Section 3.3). We perform association analysis with risk factors and aging-related morbidities, and genome-wide association study (GWAS) on the model trained on different mortality causes using the UKB data and show that ENABL Age clocks trained on different age-related outcomes can capture different aging mechanisms, which allows us to focus on specific components of aging (Section 3.4). Finally, we ensure the applicability of the ENABL Age approach to other age-related outcomes by predicting dementia using the ROSMAP dataset and validating that ENABL Age accurately captures dementia-related aging signals (Supplementary Appendix Section 1). Our ENABL Age clocks are available on an interactive website (https://suinleelab.github.io/ENABLAge) which allows users to calculate and interpret their own ENABL Age. The code for our study is available at https://github.com/suinleelab/ENABLAge.

## 2 Methods

### 2.1 Data sources and study population

This study focuses on the UK Biobank^1^ (UKB) and the National Health and Nutrition Examination Survey (NHANES)^2^ datasets. UKB participants were enrolled between 2007-2014 from 21 assessment centers across England, Wales, and Scotland. Our study includes all measurements taken during their initial visit, available on December 13th, 2021. We exclude (1) features that are missing in more than 80% of the samples, and (2) highly correlated features with correlations greater than 0.98. After excluding features, our UKB dataset has 825 features from numerous categories: demographics, blood assays, health and medical history, lifestyle and environment, physical measures, etc. The samples from two Scottish centers (Edinburgh and Glasgow) are left out as the geographical validation set (n=35,735). We predict all-cause mortality and six cause-specific mortality categories based on ICD10 codes, as inGanna and Ingelsson [15]: neoplasm, C00–D48; diseases of the circulatory system, I05–I89; diseases of the respiratory system, J09–J99; diseases of the digestive system, K20–K93; external causes of mortality and morbidity, V01–Y84; and other diseases, all remaining ICD10 codes. We remove deceased samples due to external causes of mortality and morbidity. After excluding samples, our UKB dataset has 501,366 samples aged 40-70.

The NHANES data are collected from individuals in the US between 1999-2014. We include demographic, laboratory, examination, and questionnaire features that could be automatically matched across different NHANES cycles. We exclude features that are missing for more than 50% of the participants and highly correlated features with correlations greater than 0.98. After excluding features, 158 features are left. We predict all-cause mortality and 9 cause-specific mortality categories included in the public-use mortality file: heart disease, malignant neoplasms, chronic lower respiratory disease, unintentional injuries, cerebrovascular diseases, Alzheimer’s disease, diabetes, pneumonia and influenza, and kidney disease. All remaining deaths are categorized into other causes. We remove deceased samples of unintentional injuries. After excluding samples, 47,084 samples aged 18-80+ remain in our NHANES dataset. Demographic characteristics and sample size of the data for different tasks are shown in Table 1.

**Table 1:**
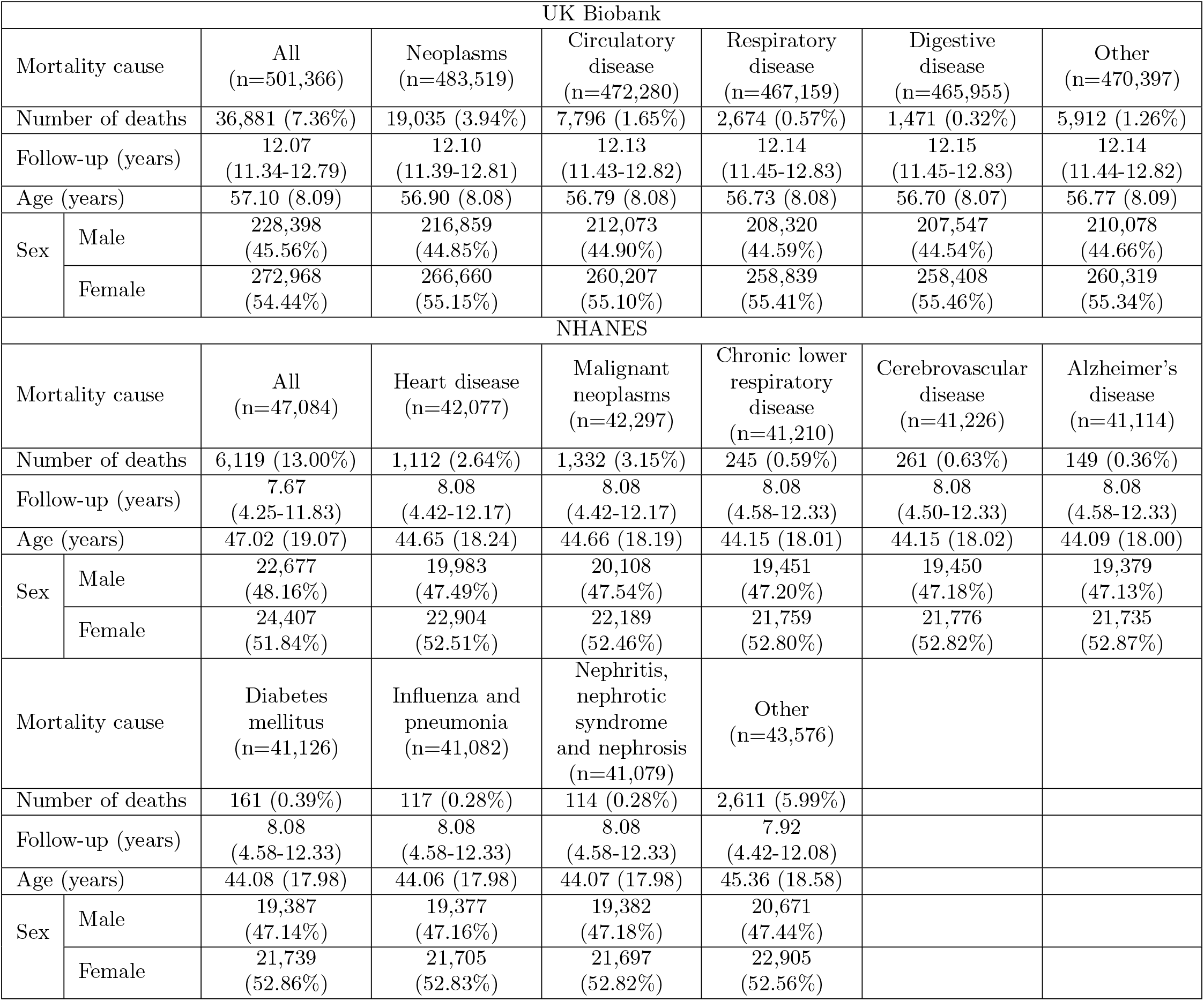
Population characteristics for the study cohorts. Data are mean(SD), median (IQR), or n/N (%).

### 2.2 Overview of the ENABL Age framework

We construct ENABL Age clocks in two stages (Figure 1A). First, we develop predictors of all-cause mortality and cause-specific mortality on all or a subset of the variables using Cox proportional hazard (CoxPH) models. When predicting cause-specific mortality, the deceased samples of all other causes are excluded. To achieve high accuracy, we use gradient boosted trees (GBTs), i.e., nonparametric models composed of iteratively trained decision trees. The final ensemble of trees can capture non-linear and interaction effects between predictors. The hyperparameters are chosen by GridSearch and 5-fold cross-validation.

**Figure 1:**
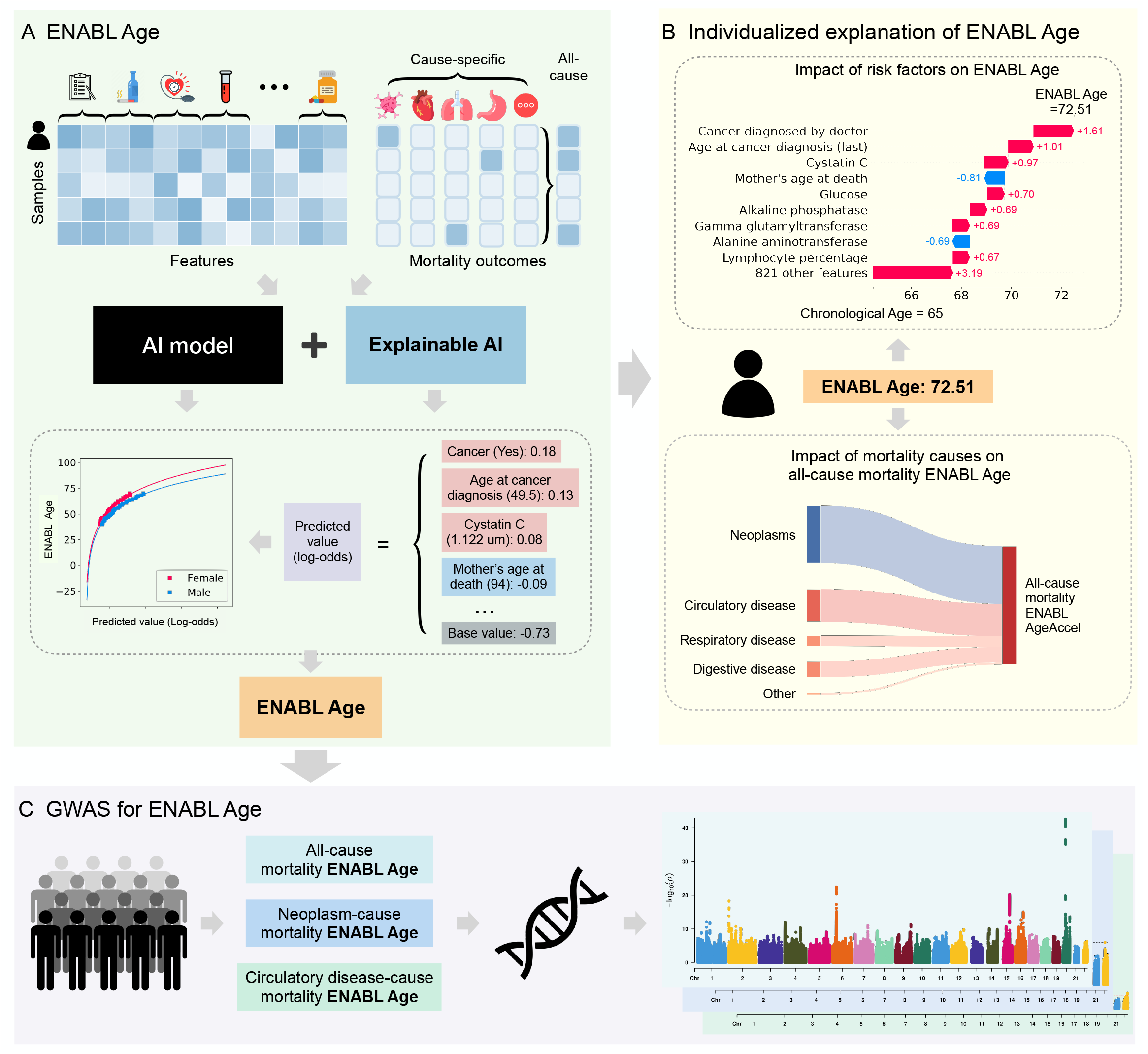
Overview of the ENABL Age framework. (A) ENABL Age directly predicts an age-related outcome of interest (e.g., all-cause mortality or cause-specific mortality) using GBTs and then nonlinearly rescales the predictions to be in units of age. (B) We extend existing explainable ML methods to calculate the contributions of ENABL Age input features in units of years, which makes it more human-interpretable. (C) We perform association analysis and GWAS on ENABL Age clocks trained on different mortality causes using the UK Biobank data to show that those trained on different age-related outcomes can capture different aging mechanisms.

Second, we fit an exponential curve to the training samples’ chronological ages and GBT model predictions (Figure 2C,F). We use the inverse function of the exponential curve to calculate our ENABL Age given a prediction (Figure 2D,G). Next, we define ENABL Age acceleration (AgeAccel) as the difference between the ENABL Age and chronological age. Finally, to obtain the contribution of different mortality causes to all-cause ENABL Age, we train GBT models to predict the all-cause ENABL Age using chronological age and cause-specific ENABL Ages for different sexes.

**Figure 2:**
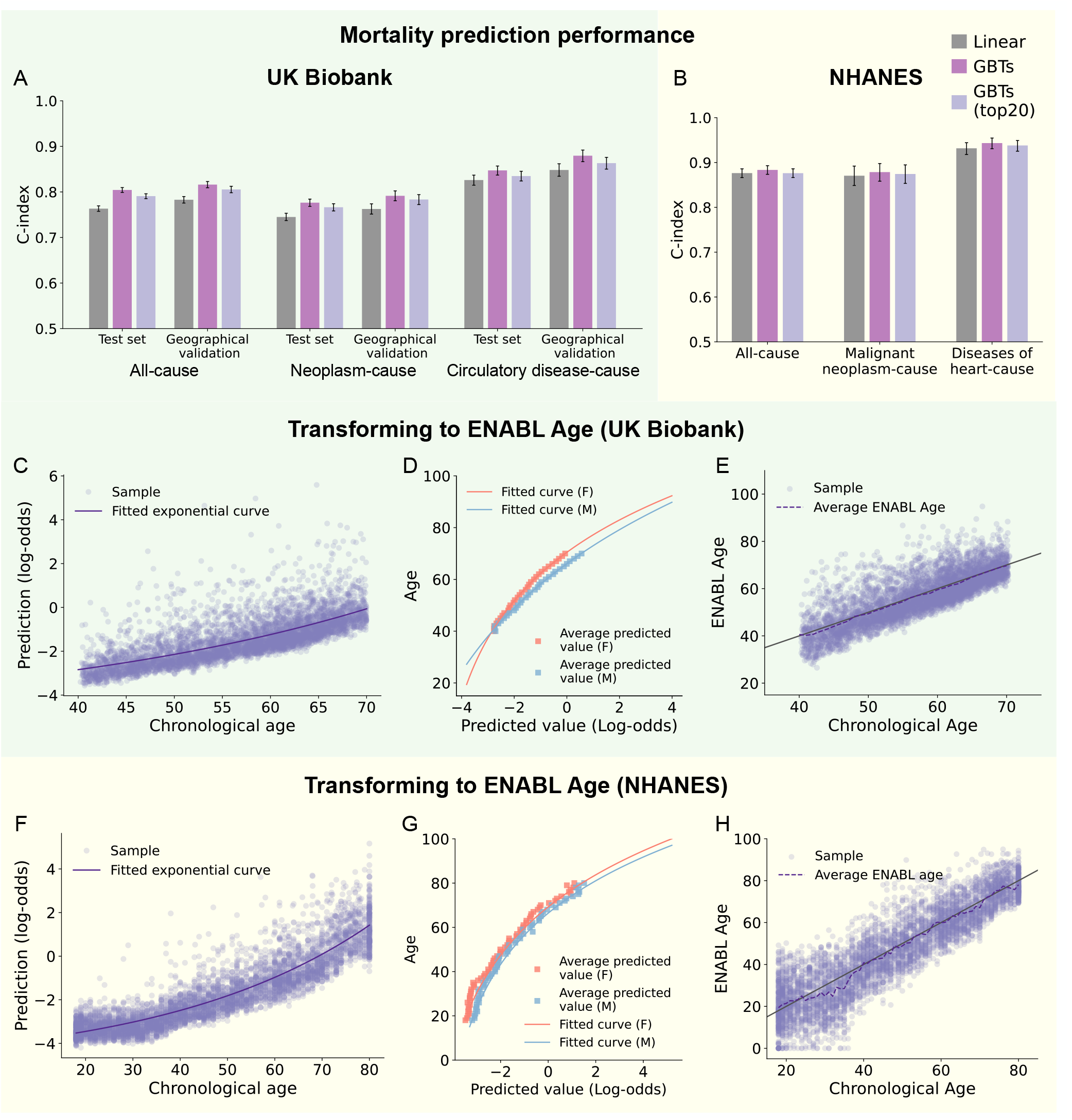
ENABL Age calculation. (A-B) The C-index of the all-cause mortality prediction and causespecific mortality prediction on UK Biobank test set, a left-out geographical validation set (more details in Section 2.1) and NHANES test set using linear Cox regression models, GBTs Cox regression models, and GBTs Cox regression models with the top 20 important features selected by a recursive feature elimination method (more details in Section Methods 2.2). Confidence intervals are computed using bootstrap resampling over the tested time points while measuring the difference of the C-index between the linear models and the GBTs with n = 1000 independently resampling. (C-D) The transformation of mortality prediction to ENABL Age on the UK Biobank dataset. (C) The scatter plot of GBTs’ predicted log-odds versus chronological ages and the fitted exponential curve on the UK Biobank dataset. (D) The curves that transform GBTs’ predicted value to ENABL Age on the UK Biobank dataset. (E) The scatter plot of ENABL Ages versus chronological ages on the NHANES dataset. (F-H) The transformation of mortality prediction to ENABL Age on the NHANES dataset. (F) The scatter plot of GBTs’ predicted log-odds versus chronological ages and the fitted exponential curve on the NHANES dataset. (G) The curves that transform GBTs’ predicted value to ENABL Age on the NHANES dataset. (H) The scatter plot of ENABL Ages versus chronological ages on the NHANES dataset.

### 2.3 ENABL Age interpretability

To identify the contribution of each feature or cause to ENABL Age (Figure 1B), we apply TreeExplainer [33]; this method provides a local (i.e., for each subject) explanation of the impact of input features on individual predictions for GBT models. TreeExplainer calculates exact SHAP [32] (SHapley Additive exPlanations) values for GBT models, which guarantee a set of desirable theoretical properties. SHAP values are *additive*; they sum to the model’s output, i.e., the log-odds for GBTs. They are also *consistent*, which means that unambiguously more important features are guaranteed to have a higher SHAP value.

To identify an individual’s strong risk factors compared to people of the same age and sex, we use the testing samples with a specific age as the explicands (i.e., samples being explained) and training samples with the same age as the baselines (i.e., samples being compared to) when calculating the SHAP values. Then, we rescale the SHAP values to the ENABL Age space based on the generalized rescale rule proposed in our previous work [6]. The rescaled SHAP values are in units of years and sum to the ENABL AgeAccel. They explicitly show the contribution of each feature to the ENABL Age. Likewise, we can calculate the SHAP values of different mortality causes to all-cause mortality ENABL Age; we do this by using the explicands and baselines of the same age. Afterwards, we rescale the SHAP values of the features to cause-specific mortality predictions to sum to the SHAP values of different mortality causes to all-cause mortality. Consequently, ENABL Age framework provides the contributions of the features to different mortality causes as well as mortality causes to all-cause mortality.

### 2.4 Statistical analysis

We impute missing values using MissForest [45]. The datasets are randomly divided into training (80%) and testing (20%) sets. The performance of the CoxPH models is measured using the concordance index (Cindex). The performance of the all-cause mortality ENABL Age clock is measured using *R*^2^. We plot KaplanMeier curves for samples with the highest 25% ENABL AgeAccel versus the lowest 25% to evaluate whether ENABL Age accurately reflect individual health status. Furthermore, to directly compare the mortality prediction power of ENABL Age with chronological age, PhenoAge [26] and BioAge [25], we train 5and 10-year mortality prediction models using different biological age accelerations adjusted for chronological age and sex, and we compare the area under the receiver operating characteristic curves (AUROCs).

We also examine the associations of ENABL AgeAccels, PhenoAgeAccel, and BioAgeAccel with a range of risk factors and age-related morbidity outcomes using UKB geographical validation data. The risk factors and outcomes are regressed separately on each of the biological age accelerations, adjusting for chronological age and sex using ordinary least squares regression (pack years of smoking, grip strength, forced expiratory volume in 1-second, waist-hip ratio), logistic regression (walking pace), and Cox regression (cancer, myocardial infarction, stroke, COPD, asthma, all-cause dementia and end-stage renal disease diagnosis), as appropriate. We report the change (i.e., beta for ordinary least squares squares regression; odds ratio for logistic regression; and hazard ratio for Cox regression) in each of the outcome measures associated with a standard unit increase in age acceleration in each of the 4 biological aging measures.

We perform GWAS on all-cause, neoplasm-cause and circulatory disease-cause mortality ENABL AgeAeecl using UKB data to validate if they can capture different aging mechanisms when trained on different tasks. After quality control, 382,152 unrelated participants and 12,755,286 SNPs in UKB data are included in our analyses. The association between ENABL AgeAccel with each SNP is examined using an efficient Bayesian linear mixed effects model, BOLT-LMM [29]. SNP p-values smaller than 5*×*10^8^ are deemed to be statistically significant.

We further perform a step-wise model selection procedure on the genome-wide SNP summary statistics to identify independent signals (*p<* 5 *×* 10^8^) using the Conditional and Joint association analysis (COJO) algorithm. The loci marked by the selected SNPs were mapped to genes based on GRCh37/hg19 coordinates and are used in searches for published GWAS associations based on the GWAS catalog [4]. We also calculate the genetic correlations of ENABL AgeAccels with several health-related traits, including anthropometric traits (e.g., BMI), adiposity (e.g., whole body/leg/arm/trunk fat mass), longevity (mother/father’s longevity), lifestyle (e.g., smoking) and several diseases (e.g., heart attack, heart failure, angina, stoke and hypertension) by LD score regression [3] using GWAS summary statistics, which are downloaded from http://www.nealelab.is/uk-biobank. More details of our methods can be found in Supplementary Methods.

## 3 Results

### 3.1 ENABL Age clocks capture non-linear relationships

Linear models are commonly used in biological age studies because they are easy to interpret – if a feature is assigned a positive weight, then a higher value of that feature corresponds to a higher predicted age, and the more positive the weight, the stronger the positive effect of that feature [42]. However, more expressive models, such as tree-based ones, can achieve higher predictive accuracy across many datasets by learning non-linear relationships between features and the outcome variable. Gradient boosted trees (GBTs) have achieved state-of-the-art performance in many domains [10, 48, 52], especially on mortality prediction tasks [33, 40]. In our study, GBT models also outperform linear models across almost all mortality prediction tasks we consider (Figures 2A,B; Supplementary Figure 2). The superior prediction performance of tree models indicates that they can effectively capture signals relevant to mortality, which are also strongly related to aging.

Using non-linear models to build biological age also benefits from minimal assumptions about the data distribution. Linear models make several assumptions (e.g., linearity, independence, normality, etc.) that constrain the relationships we can learn and the features we can use. These assumptions make their use problematic to apply to datasets with large numbers of features or highly correlated features [42]. To accommodate these assumptions, it is common to perform complicated feature selection and data transformations prior to fitting linear models. In comparison, tree-based models make minimal assumptions about the data distribution and need no data transformations.

Unlike GrimAge [31], which maps mortality predictions to aging using linear transformation, we use an exponential curve to fit mortality predictions (i.e., log-odds of GBT models) and chronological ages (Figures 2 C,F). Figures 2D,G show that the inverse functions of the exponential curves accurately model the relationship between the average predicted values and chronological ages. As shown in Figures 2E,H, ENABL Age is highly correlated with chronological age (Pearson’s *r* =0.79). In Supplementary Figures 3, we also show that the ENABL Age of currently living samples at mortality data collection time is consistently higher than those of deceased samples, illustrating the effectiveness of our model.

### 3.2 ENABL Age is a strong indicator of individual health status

Figure 3 shows Kaplan-Meier survival curves of all-cause and neoplasm-cause mortality for UKB and NHANES samples, stratified into the healthy (lowest 25%) and unhealthy (highest 25%) age groups according to ENABL AgeAccels. We find that those with the highest ENABL AgeAccels relative to their chronological ages have much steeper declines in survival over the 14 (UKB) and 16 (NHANES) years of follow-up. For neoplasm-cause mortality, which has a low base rate (3.39% for UKB, 3.15% for NHANES), the difference in survival probability between healthy and unhealthy aging subjects is especially prominent. Altogether, these results suggest that ENABL Age clock successfully stratifies unhealthy and healthy individuals, which in turn indicates that ENABL Age accurately reveals individual’s health status.

**Figure 3:**
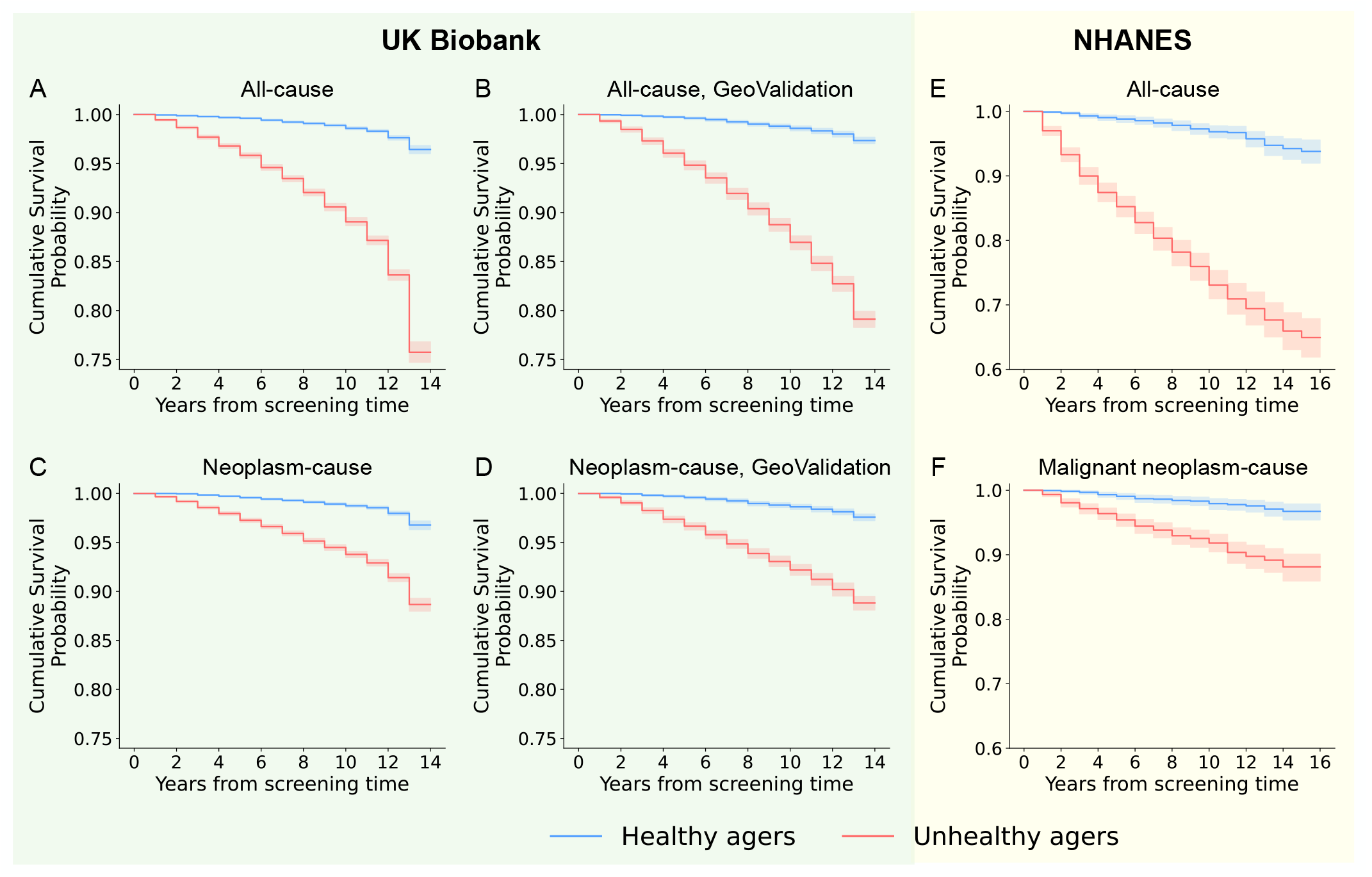
ENABL Age clocks successfully stratify unhealthy and healthy individuals. (A-B) Kaplan–Meier curves for persons in the highest 25% (unhealthy agers) versus the lowest 25% (healthy agers) of all-cause mortality ENABL AgeAccel in the UK Biobank test set and geographical validation set. (C-D) Kaplan–Meier curves for persons in the highest 25% (unhealthy agers) versus the lowest 25% (healthy agers) of neoplasm-cause mortality ENABL AgeAccel in the UK Biobank test set and geographical validation set. (E-F) Kaplan–Meier curves for persons in the highest 25% (unhealthy agers) versus the lowest 25% (healthy agers) of all-cause mortality ENABL AgeAccel and malignant neoplasm-cause mortality ENABL AgeAccel in NHANES test set.

Then, we directly evaluate the mortality prediction power of ENABL AgeAccel, PhenoAgeAccel and BioAgeAccel by training 5- and 10-year all-cause mortality prediction models adjusted by chronological age and sex. PhenoAge [26] and BioAge [25] are two second-generation biological age clocks that are also built with phenotypic features; it is worth mentioning that PhenoAge also uses all-cause mortality information during training. We calculate the PhenoAge and BioAge using the formulae in their original papers. To show the effectiveness of ENABL Age framework, we also train all-cause mortality ENABL Age clocks using the features included in PhenoAge and BioAge. We further build all-cause mortality ENABL Age clocks using other subsets of features: laboratory features in the four most popular blood panels (ENABL Age-L) – CBC (Complete Blood Count), CMP (Comprehensive Metabolic Panel), LP(Lipoprotein (a)) and WBC (White Blood Cell Count), the top 20 most important questionnaire features (ENABL Age-Q), and the top 20 most important features (ENABL Age-20).

Figures 4A-F show the AUROCs of the 5- and 10-year mortality prediction models using different ages and feature subsets. We see that using the same features, ENABL Age clocks achieve higher AUROCs than PhenoAge and Bioage, indicating that it is a stronger predictor of mortality. Importantly, both PhenoAge and BioAge use C-reactive protein as an input, which is not collected in the most popular blood panels. ENABL Age-L has similar mortality prediction power to ENABL Age clock using the features that PhenoAge and BioAge rely on, suggesting that C-reactive protein can be replaced by features collected in the most popular blood panels. ENABL Age-Q and ENABL Age-20 achieve performance similar with ENABL Age clock using all the features, showcasing the effectiveness of the selected features. In particular, ENABL Age-L and ENABL Age-Q are both low-cost and highly accurate; ENABL Age-L relies only on popular blood tests and is usable by medical professionals, whereas ENABL Age-Q depends only on questionnaire features and can be used by non-professional healthcare consumers.

**Figure 4:**
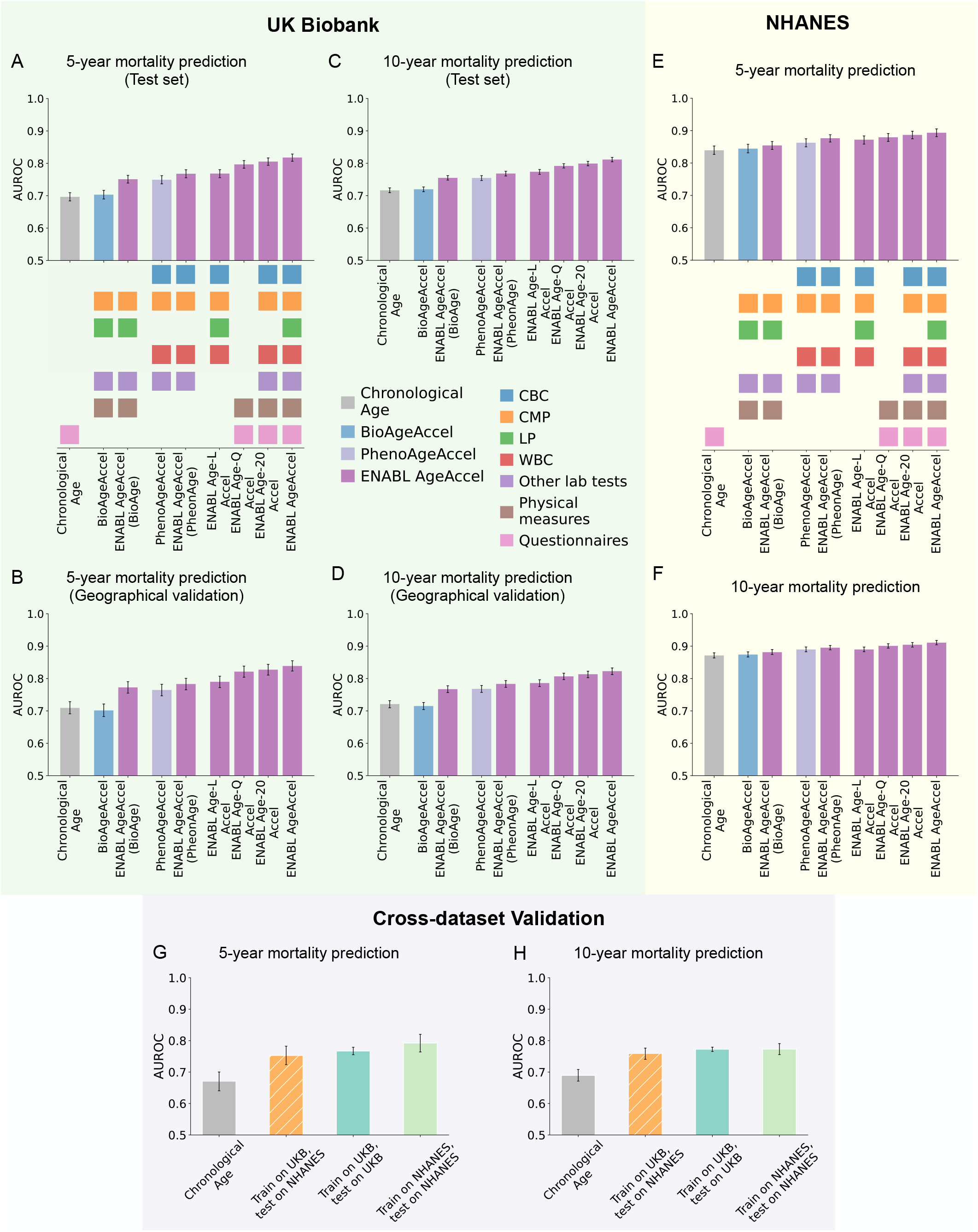
ENABL AgeAccels have high mortality prediction power. (A-D) The AUROCs of the 5-year and 10-year mortality prediction GBT models of ENABL AgeAccels, PhenoAgeAccel and BioAgeAccel adjusted by chronological age and sex trained and test on the UKB dataset. We also validate the generalizability of ENABL Age clocks on the UKB geographical validation set. “Chronological Age” refers to the GBT models trained by chronological age and sex. “BioAge” and “PhenoAge” are calculated using the formulae in their original papers. “ENABL Age (BioAge)” and “ENABL Age (PhenoAge)” are calculated by the ENABL Age framework using only the features included in BioAge and PhenoAge, respectively. “ENABL Age-L” refers to the ENABL Age clock that uses laboratory features in the four most popular blood panels – CBC, CMP, LP and WBC. “ENABL Age-Q” refers to the ENABL Age clock that uses the top 20 most important questionnaire features selected by recursive feature elimination. “ENABL Age-20” refers to the ENABL Age clock that uses the top 20 most important features out of all possible features selected by recursive feature elimination (Supplementary Methods 2.2; Supplementary Figures 2B,D). Colored squares indicate the feature types/laboratory panels (CBC – Complete Blood Count), CMP – Comprehensive Metabolic Panel, LP –Lipoprotein (a) and WBC – White Blood Cell Count) used to calculate biological ages. (E-F) The AUROCs of the 5-year and 10-year mortality prediction GBT models of ENABL AgeAccels, PhenoAgeAccel and BioAgeAccel adjusted by chronological age and sex trained and tested on the NHANES dataset. (G-H) The cross-dataset validation of the 5- and 10-year mortality prediction models using ENABL Age-L. “Chronological Age” refers to the mortality prediction model using chronological age and sex trained and tested on NHANES samples aged 40-70. “Train on UKB, test on NHANES” refers to cross-dataset validation of ENABL Age clock. Specifically, we train ENABL Age-L on the UKB samples using the features in the four popular blood panels that overlap in the UKB and NHANES datasets and use it to evaluate the ENABL Age-L for NHANES samples. Then, we train and test the mortality prediction models using chronological age, sex and ENABL Age-L acceleration (trained on UKB samples) on NHANES samples aged 40-70. “Train on UKB, test on UKB” and “Train on NHANES, test on NHANES” refers to the mortality prediction model using chronological age, sex and ENABL Age-L acceleration trained and evaluated on UKB samples or NHANES samples aged 40-70, respectively.

Figures 4B,D show that ENABL Age clock successfully generalizes to a geographically distinct UKB validation set (i.e., samples collected in two Scottish centers). We also perform a cross-dataset validation using ENABL Age-L by training it on UKB samples using the features in the four popular blood panels that overlap between UKB and NHANES datasets and evaluating the 5- and 10-year mortality prediction power on NHANES testing samples aged 40-70. From Figures 4G,H, we see that the AUROC of the ENABL Age clock trained on UKB and tested on NHANES is similar to the AUROCs of the ENABL Age clocks trained and tested within the same dataset, confirming the external generalizability of ENABL Age clock.

Given that biological age should reflect individuals’ health status, we examine whether ENABL Age clocks relate to diverse risk factors and age-related morbidity outcomes. We examine the associations of ENABL AgeAccels, PhenoAgeAccel, BioAgeAccel with a range of risk factors (pack years of smoking, walking pace, grip strength, forced expiratory volume in 1-second and waist-hip ratio) and age-related morbidity outcomes (cancer, myocardial infarction, stroke, COPD, asthma, dementia and end-stage renal disease) on the UKB geographical validation data. Figure 5 shows the change in each risk factor and age-related outcome associated with a one-year increase in biological age acceleration while adjusting for chronological age and sex. We observe strong and significant associations between ENABL AgeAccels and all risk factors and age-related outcomes we consider; the associations are stronger than PhenoAgeAccel and BioAgeAccel for most of the risk factors and age-related outcomes. Although BioAgeAccel has stronger associations with waist-hip ratio and myocardial infarction than ENABL AgeAccels, it shows no significant associations with 4/7 of the age-related morbidity outcomes we consider (i.e., cancer, COPD, asthma, and all-cause dementia).

**Figure 5:**
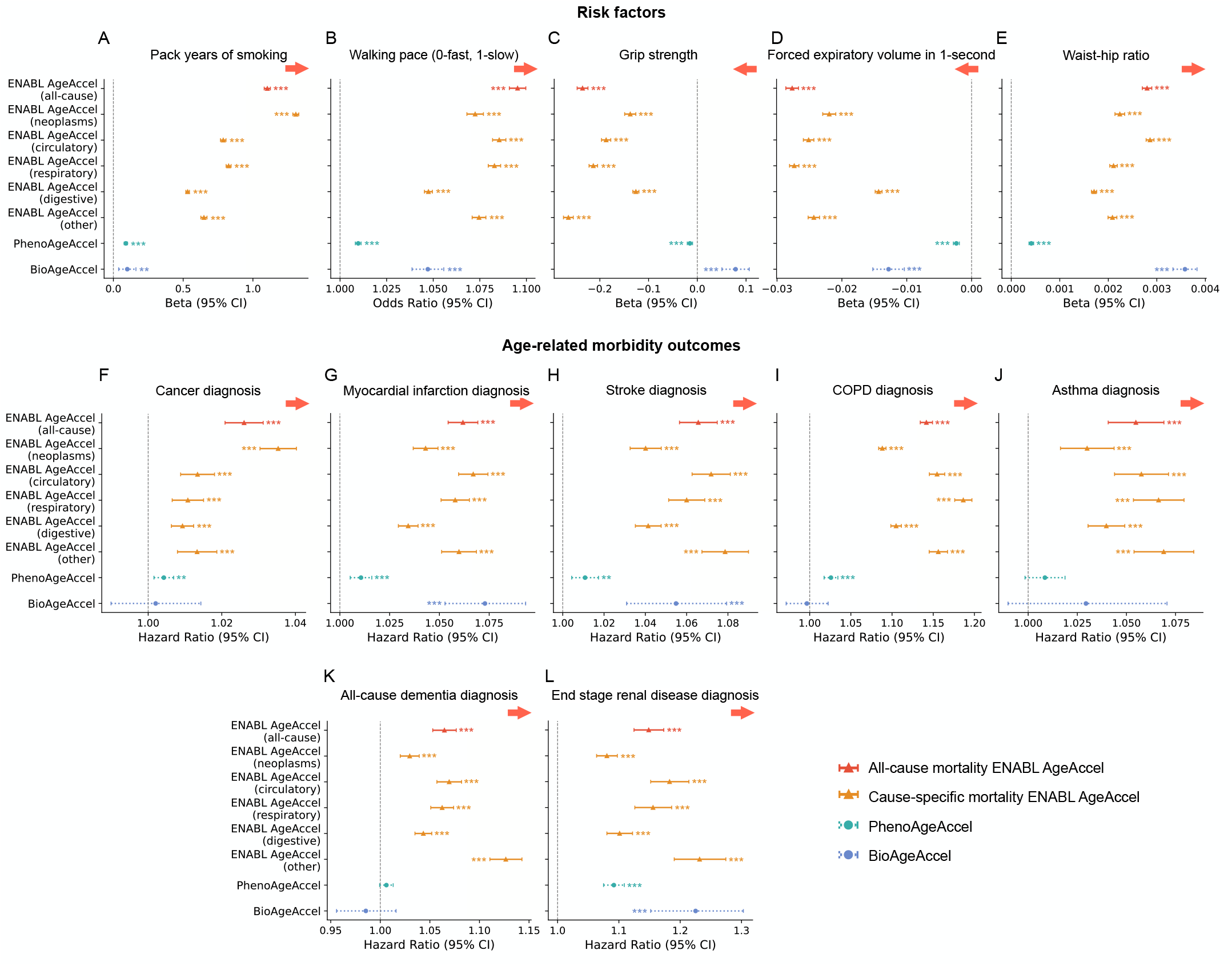
ENABL AgeAccels are strongly associated with diverse risk factors and age-related morbidity outcomes. (A-L) Associations of age accelerations (ENABL AgeAccel, PhenoAgeAccel, BioAgeAccel) with phenotypes (i.e., pack years of smoking, walking pace, grip strength, forced expiratory volume in 1-second, waist-hip ratio, cancer, myocardial infarction, stroke, COPD, asthma, dementia and end-stage renal disease diagnosis; Supplementary Methods 4). The risk factors and outcomes are regressed separately on each of the biological age accelerations, adjusting for chronological age and sex using ordinary least squares regression (pack years of smoking, grip strength, forced expiratory volume in 1-second, waist-hip ratio), logistic regression (walking pace), and Cox regression (cancer, myocardial infarction, stroke, COPD, asthma, all-cause dementia and end-stage renal disease diagnosis), as appropriate.

As an approach that estimates the second-generation biological age, ENABL Age framework incorporates age-related outcomes by directly predicting them, so that it powerfully and robustly captures aging morbidity and mortality information. Further, ENABL Age framework can use any available features; thus, it can capture more comprehensive aging mechanisms than PhenoAge and BioAge, which use only a small number of features correlated with aging or mortality. In summary, ENABL Age clocks show strong and robust performance in identifying unhealthy individuals and predicting mortality. Moreover, they have strong associations with a variety of meaningful risk factors and age-related morbidity outcomes.

### 3.3 ENABL Age is interpretable and exposes individualized aging explanations

ENABL Age clock makes mortality risk scores easier to understand. Mortality risk scores can be hard for a general audience to understand because their scale (e.g., predicted probability, log-odds) is not inherently meaningful. To fix this, our ENABL Age clock produces estimates of an individual’s age as a function of their mortality risk. For example, an ENABL Age of 65 means that this individual’s risk score is most similar to the risk scores of people who have a chronological age of 65. In doing so, we provide a more accessible estimate of mortality risk contextualized by chronological age.

Individualized explanations of biological age (i.e., the individualized contributions of features to biological age) are important for understanding complex aging mechanisms on a personal basis and further guiding clinical decision-making. We make ENABL Age interpretable by calculating SHAP values for GBT models and rescaling them into units of years. To do so, we first calculate SHAP values for an explicand (i.e., a sample being explained) relative to baselines (i.e., samples being compared to) with the same age and sex as the explicand in order to identify risk factors while controlling for age and sex. We then rescale the SHAP values to be in units of years based on the generalized rescale rule proposed in our previous work [6].

Figures 6A,C show rescaled SHAP values of the top 10 most important features for all-cause mortality and neoplasm-cause ENABL Age for female samples aged 65. Figures 6B,D show an individualized explanation for a 65-year-old woman’s all-cause mortality and neoplasm-cause mortality ENABL Age. We observe that the individual’s all-cause and neoplasm-cause mortality ENABL Age are 72.94 and 73.82, respectively, higher than their chronological age, i.e., 65. Based on the SHAP values, features that increase ENABL Ages include age at cancer diagnosis, sex hormone binding globulin, long-standing illness, disability or infirmity; features that decrease ENABL Ages include past tobacco smoking (not smoking) and type of cancer tumour (epithelial). This individual’s ENABL Ages are higher than the average age of the baselines (approximate to chronological age), so the features that increase them outweigh those that decrease them.

**Figure 6:**
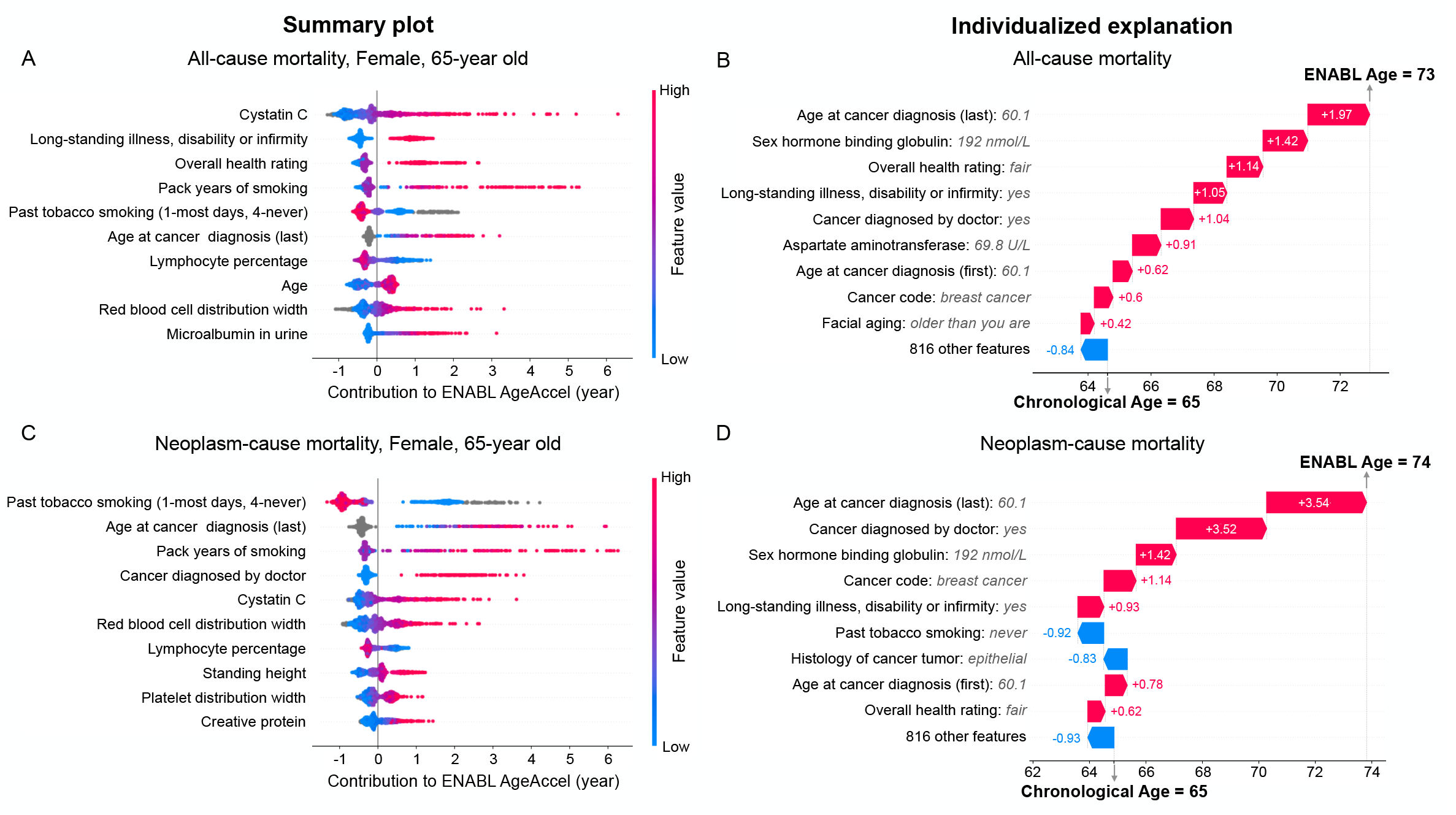
ENABL Age exposes individualized aging explanations. (A,C) SHAP summary plot for ENABL Age clocks trained on all-cause mortality and neoplasm-cause mortality using UKB datasets for female samples aged 65-years old. The plot shows the most impactful features on ENABL Age (ranked from most to least important). It also shows distribution of the impacts of each feature on ENABL AgeAccel, which includes a set of plots where each dot corresponds to an individual. The colors represent feature values for numeric features: red for larger values, blue for smaller, and grey for not applicable (e.g., currently smoke on most or all days for past tobacco smoking). The thickness of the line (which actually comprises many individual dots) is determined by the number of examples at a given value, where dots spread out vertically if there are many examples. A negative rescaled SHAP value (extending to the left) indicates reduced ENABL AgeAccel, while a positive one (extending to the right) indicates increased ENABL AgeAccel. The SHAP values are calculated using explicands and baselines that have the same age and sex (i.e., females aged 65-years old). Compared with Supplementary Figures 4A,B which use the explicands and baselines from the general polulation, age and sex are no longer the most important features, and strong risk factors for a specific age and sex are revealed. (B,D) The individualized explanation of all-cause mortality ENABL Age and neoplasm-cause mortality ENABL Age for a single female aged 65-year old. The output value (the gray dashed line with the number at the top of the plot) shows ENABL Age for that individual. The base value (the gray dashed line with the number at the bottom of the plot) approximates chronological age (i.e., 65). The features in red increase ENABL AgeAccel, and those in blue decrease it.

To more comprehensively explain individuals’ aging processes, we can also obtain the contribution of different aging aspects (mortality causes) to all-cause mortality ENABL Age. To do so, we train models to predict the all-cause mortality ENABL Age using chronological age and cause-specific mortality ENABL Ages for different sexes (*R*^2^ = 0.9722 for female, *R*^2^ = 0.9721 for male); we then rescale the SHAP values of different mortality causes to all-cause mortality ENABL AgeAccel. Consequently, we achieve the two-layer explanations of all-cause mortality ENABL AgeAccel, i.e., the features to different mortality causes as well as mortality causes to all-cause mortality ENABL Age.

Figure 7A shows the two-layer explanations for the same individual as in Figure 6B,D. The features in red increase all-cause mortality ENABL AgeAccel, and those in blue decrease it. We see that all of the mortality causes increase all-cause mortality ENABL AgeAccel, with neoplasm making the largest contribution. Figure 7B shows the two-layer explanations of a healthier individual. Here, we see that most of the mortality causes (i.e., neoplasms, circulatory disease, and other) decrease the all-cause mortality ENABL AgeAccel and this individual has a negative all-cause mortality ENABL AgeAccel. We also observe that cystatin C and sex hormone binding globulin decrease this individual’s ENABL AgeAccel result.

**Figure 7:**
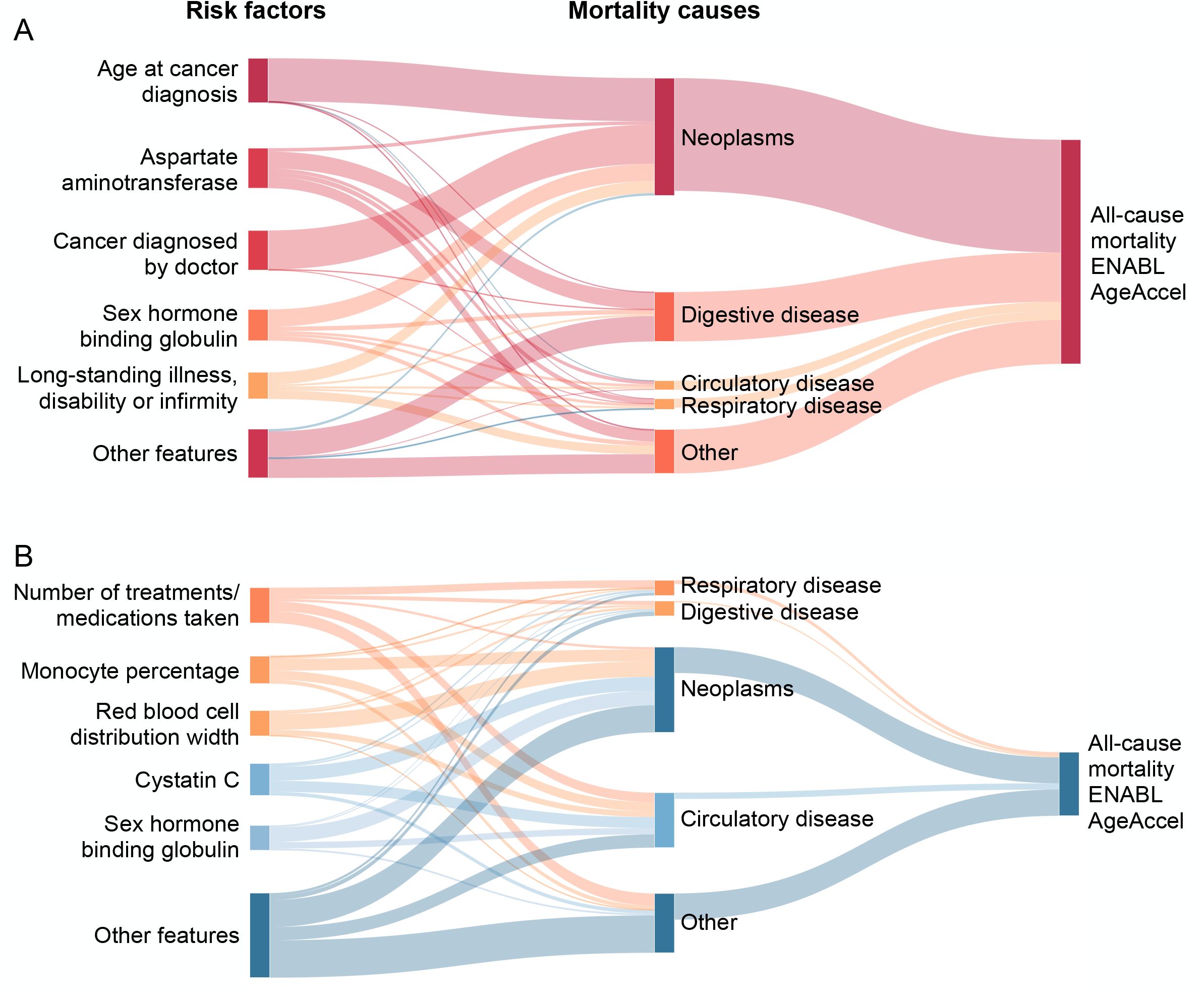
Two-layer explanations of all-cause mortality ENABL Age. (A-B) The two-layer all-cause mortality ENABL Age explanations for two samples from UK Biobank dataset. The lines in red indicate positive rescaled SHAP values that increase the ENABL AgeAccel; those in blue indicate negative rescaled SHAP values that decrease the ENABL AgeAccel.

Being able to interpret a personal ENABL Age can help individuals improve their health awareness and adjust their lifestyle accordingly. It also assists medical professionals in efforts to elucidate complex and interrelated aging mechanisms and potentially guide clinical decision-making (the examples from NHANES dataset can be found in Supplementary Figure 5).

### 3.4 ENABL Age clocks capture varied aging mechanisms

Previous studies have suggested that different biological age clocks for the same individual may be capturing distinct aspects of the aging process, so it is vital to understand what aging signals are being captured [21, 23, 42]. For ENABL Age, we directly predict the age-related outcome and map the predictions to age so we can determine what aging signal our ENABL Age clock captures by specifying the age-related outcome. In Figure 5, we see that ENABL AgeAccels trained on different mortality causes have strong association with different diseases: neoplasm-cause mortality ENABL AgeAccel has stronger association with cancer, circulatory-disease ENABL AgeAccel has stronger associations with myocardial infarction and stroke, and respiratory-disease cause mortality ENABL AgeAccel has stronger association with COPD. Together, these results suggest that each ENABL Age clock trained on specific mortality cause successfully capture appropriate disease-causing aging mechanisms.

To validate if the ENABL Age clocks trained on different tasks can capture different aging mechanisms, we perform GWAS on all-cause, neoplasm-cause and circulatory disease-cause mortality ENABL AgeAccel using UKB data. Since genes can be tied experimentally to biological aging processes, showing the different genetic architecture of ENABL Age clocks can help us understand the aging signals they capture.

We identify 93 conditionally independent associations for all-cause mortality ENABL AgeAccel, 48 for neoplasm-cause mortality ENABL AgeAccel, and 174 for circulatory disease-cause mortality ENABL AgeAccel (*p<* 5 *×* 10^−8^) (Manhattan plot in Figure 8). From the Manhattan plots, we see that the associated SNPs differ for each ENABL Age clock. The selected mapped genes of the lead SNPs are shown in Table 2. For all-cause mortality ENABL AgeAccel, the mapped genes are associated with anthropometric measures (e.g., BMI, waist-hip ratio), blood count measures (e.g., platelet count, neutrophil count), alcohol consumption, smoking behavior and different cancers, which are all age- and health-related traits [42]. For neoplasm-cause mortality ENABL AgeAccel, the mapped genes are associated with different cancers (e.g., breast cancer, lung cancer), smoking behavior, BMI and blood count measures. For circulatory disease-cause mortality ENABL AgeAccel, the mapped genes are associated with circulatory diseases (e.g., coronary artery disease, myocardial infarction) and protein levels. Previous work also shows that protein levels are associated with cardiovascular disease [50, 38]. In Table 2, we see that the genes identified by ENABL AgeAccels are also identified to be associated with other biological ages, multivariate aging traits and longevity in previous work [34, 23], suggesting that our ENABL Age clocks concur with and potentially complement prior biological age clocks.

**Figure 8:**
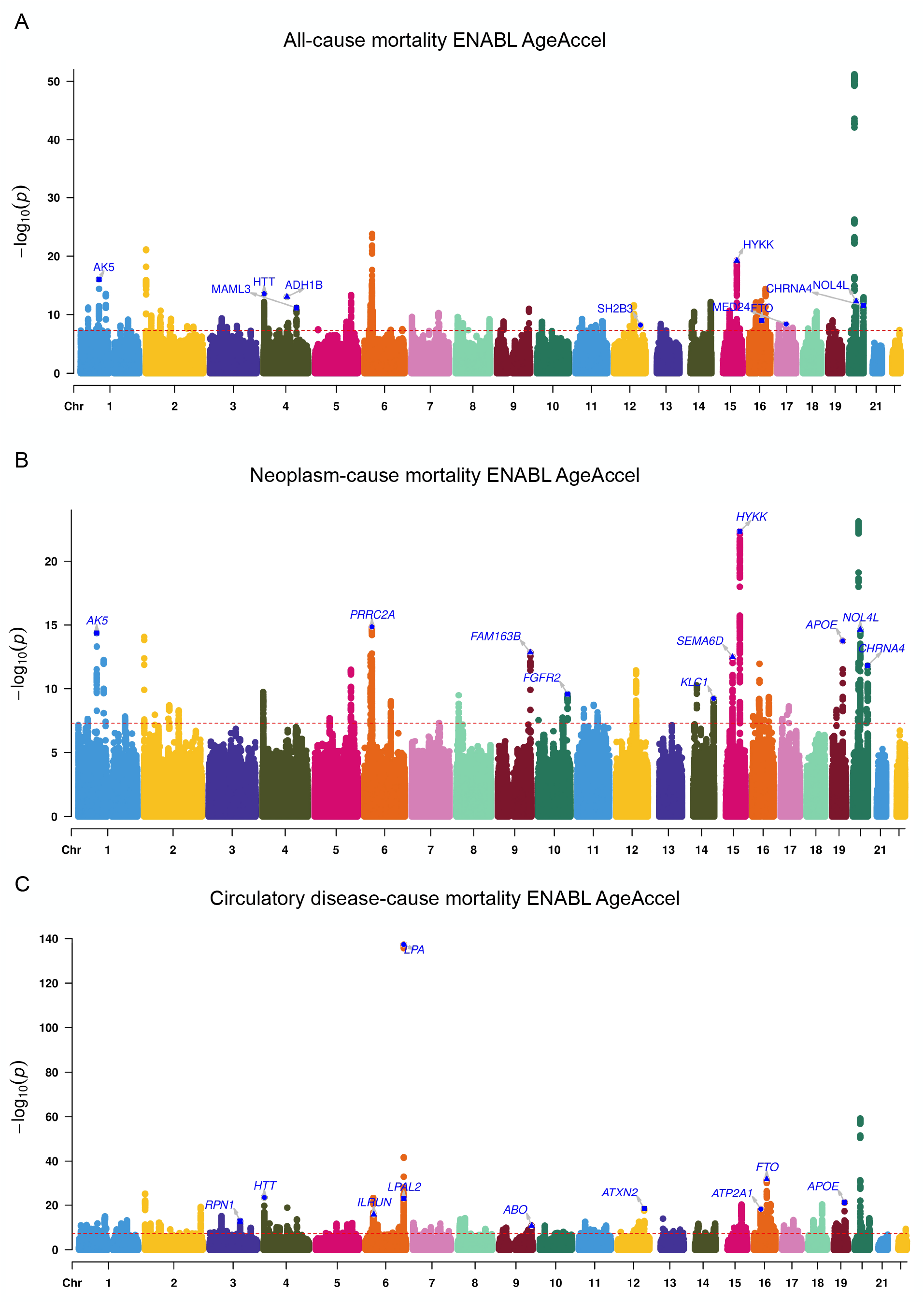
Manhattan plots for all-cause mortality ENABL AgeAccel, neoplasm-cause mortality ENABL AgeAccel and circulatory disease-cause mortality ENABL AgeAccel (colors to separate adjacent chromosomes without other indications).

**Table 2:** The selected genetic loci that are significantly associated with ENABL AgeAccel for all-cause mortality, neoplasm-cause mortality, and circulatory disease-cause mortality. The loci are mapped to interesting genes associated with health-related traits.

| SNP | Chr | bJ | pJ | Gene | Reported trait(s) | Other aging trait(s) |
| --- | --- | --- | --- | --- | --- | --- |
| **All-cause mortality ENABL AgeAccel** |  |  |  |  |  |  |
| rs71658797 | 1 | 0.18 | 1.29E-16 | AK5 | BMI, Estimated glomerular filtration rate, Lung cancer |  |
| rs362307 | 4 | 0.18 | 3.63E-11 | HTT | Type 2 diabetes, Mean platelet volume, Waist-hip ratio, Neutrophil count | Parental longevity, Multivariate aging |
| rs1229984 | 4 | 0.34 | 3.86E-13 | ADH1B | Alcohol consumption, Protein quantitative trait loci (liver), Esophageal cancer, BMI |  |
| rs809955 | 4 | -0.10 | 5.41E-12 | MAML3 | BMI, Smoking initiation, Type 2 diabetes, C-reactive protein levels |  |
| rs3184504 | 12 | -0.08 | 6.59E-09 | SH2B3 | Platelet count, Blood pressure, Hemoglobin, Monocyte count, Colorectal cancer, Cystatin C levels, Coronary artery disease | Longevity, Parental longevity |
| rs9788721 | 15 | -0.13 | 1.15E-19 | HYKK | Post bronchodilator FEV1/FVC ratio, Lung cancer, Smoking behaviour, COPD | Parental longevity, Multivariate aging |
| rs7206629 | 16 | 0.09 | 6.90E-10 | FTO | BMI, Type 2 diabetes, Obesity, Breast cancer, High density lipoprotein cholesterol levels | PhenoAgeAccel, Longevity |
| rs34003767 | 17 | -0.09 | 4.40E-09 | MED24 | Asthma and cardiovascular disease, White blood cell count | GrimAgeAccel |
| rs159428 | 20 | 0.10 | 2.60E-12 | NOL4L | Monocyte count, Smoking status, Red cell distribution width, Mean corpuscular hemoglobin | Multivariate aging |
| rs4809542 | 20 | 0.19 | 1.12E-11 | CHRNA4 | Smoking behaviour, COPD, Aerodigestive squamous cell cancer | Parental longevity |
| **Neoplasm-cause mortality ENABL AgeAccel** |  |  |  |  |  |  |
| rs71658797 | 1 | 0.17 | 7.33E-15 | AK5 | BMI, Estimated glomerular filtration rate, Lung cancer |  |
| rs10885 | 6 | 0.16 | 5.30E-17 | PRRC2A | BMI, Body fat percentage, Smoking status, Cervical cancer |  |
| rs3025316 | 9 | 0.17 | 2.45E-13 | FAM163B | Smoking cessation, Serum alkaline phosphatase levels, White blood cell count |  |
| rs2981575 | 10 | -0.09 | 2.00E-10 | FGFR2 | Breast cancer, Cancer, Gamma glutamyl transpeptidase |  |
| rs61637848 | 14 | 0.1 | 5.05E-10 | KLC1 | BMI, Urate levels, Neuropsychiatric disorders, Smoking behaviour, Breast cancer |  |
| rs9788721 | 15 | -0.15 | 4.67E-23 | HYKK | Post bronchodilator FEV1/FVC ratio, Lung cancer, Smoking behaviour, COPD, Parental longevity | Parental longevity, Multivariate aging |
| rs11638216 | 15 | -0.11 | 2.34E-13 | SEMA6D | BMI, Alcohol consumption, Smoking status, Lung cancer | Parental longevity |
| rs429358 | 19 | -0.15 | 1.64E-14 | APOE | Blood protein levels, Triglycerides, Total cholesterol levels, Alzheimer's disease | PhenoAgeAccel, BioAgeAccel, Longevity, Parental longevity, Multivariate aging |
| rs6141319 | 20 | 0.12 | 3.20E-15 | NOL4L | Monocyte count, Smoking status, Red cell distribution width, Non-melanoma skin cancer | Multivariate aging |
| rs4809542 | 20 | 0.20 | 5.19E-12 | CHRNA4 | Smoking behaviour, COPD, Aerodigestive squamous cell cancer | Parental longevity |
| **Circulatory disease-cause ENABL AgeAccel** |  |  |  |  |  |  |
| rs55683935 | 3 | -0.31 | 9.96E-14 | RPN1 | Monocyte count, Eosinophil counts, Basophil count, Neutrophil count | HorvathAgeAccel |
| rs362307 | 4 | 0.28 | 2.12E-19 | HTT | Mean platelet volume, Waist-hip ratio, Neutrophil count | Parental longevity, Multivariate aging |
| rs11803927 | 6 | 0.78 | 1.46E-150 | LPA | Lipoprotein levels, Coronary artery disease, Peripheral artery disease, Unstable angina pectoris, Myocardial infarction | Longevity, Parental longevity, Multivariate aging |
| rs14755559 | 6 | 0.90 | 6.10E-27 | LPAL2 | Lipoprotein levels, Triglyceride levels in VLDL, Coronary artery disease | Parental longevity |
| rs2744961 | 6 | 0.14 | 5.16E-16 | ILRUN | BMI, HDL cholesterol levels, Coronary artery disease, Myocardial infarction |  |
| rs550057 | 9 | 0.12 | 5.31E-11 | ABO | Blood protein levels, Cholesterol, Venous thromboembolism, Coronary artery disease, Hemoglobin concentration | PhenoAgeAccel |
| rs10774625 | 12 | -0.14 | 3.67E-19 | ATXN2 | Diastolic blood pressure, Platelet count, LDL cholesterol levels, Hemoglobin concentration, Coronary artery disease | Parental longevity, Multivariate aging |
| rs9923147 | 16 | 0.20 | 9.02E-33 | FTO | BMI, Type 2 diabetes, Obesity, High density lipoprotein cholesterol levels, C-reactive protein | PhenoAgeAccel, Longevity |
| rs7189927 | 16 | -0.15 | 5.34E-18 | ATP2A1 | BMI, Hip circumference, Type 2 diabetes, Mean corpuscular volume |  |
| rs7412 | 19 | -0.29 | 2.60E-22 | APOE | Blood protein levels, Triglycerides, Total cholesterol levels, Alzheimer's disease | PhenoAgeAccel, BioAgeAccel, Longevity, Parental longevity, Multivariate aging |

In Table 3, we show the genetic correlation between ENABL AgeAccels, other biological ages, and healthrelated traits, including anthropometric traits (e.g., BMI), adiposity (e.g., whole body/leg/arm/trunk fat mass), longevity (mother/father’s longevity), lifestyle (e.g., smoking) and several diseases (e.g., cancer, heart attack, heart failure, angina, stroke and hypertension). ENABL AgeAccels are significantly associated with all health-related traits we consider, and the associations are stronger than they are for other biological age accelerations (i.e., GrimAgeAccel, PhenoAgeAccel, Hannum AgeAccel and Horvath AgeAccel). Moreover, we see that smoking and cancer have the highest genetic correlation with the neoplasm-cause mortality ENABL AgeAccel, and circulatory diseases (e.g., heart attack, heart failure, stroke) have the highest genetic correlation with the circulatory-cause mortality ENABL AgeAccel. Overall, the GWAS and genetic correlation results highlight the different genetic architecture of different ENABL AgeAccels, which confirm that our clocks trained on different age-related tasks capture meaningful aging mechanisms.

**Table 3:** The genetic correlations between ENABL AgeAccels, other biological age accelerations, and healthrelated traits, including anthropometric traits (e.g., BMI), adiposity (e.g., whole body/leg/arm/trunk fat mass), longevity (mother/father’s longevity), lifestyle (e.g., smoking) and several diseases (e.g., heart attack, heart failure, angina, stoke and hypertension). The genetic correlations of GrimAgeAccel, PhenoAgeAccel, Hannum AgeAccel and Horvath AgeAccel are reported in McCartney et al. [34].

| Trait | All-cause<br>mortality<br>ENABL<br>AgeAccel | Neoplasm-cause<br>mortality<br>ENABL<br>AgeAccel | Circulatory<br>disease-cause<br>mortality<br>ENABL<br>AgeAccel | GrimAgeAccel | PhenoAgeAccel | Hannum<br>AgeAccel | Horvath<br>AgeAccel |
| --- | --- | --- | --- | --- | --- | --- | --- |
| BMI | 0.6216*** | 0.5426*** | **0.6611\*\*\*** | 0.3683*** | 0.2311*** | 0.1114** | 0.0637 |
| Whole body fat mass | 0.6184*** | 0.5993*** | **0.6194\*\*\*** | 0.3362*** | 0.2326*** | 0.0912* | 0.0647 |
| Leg fat mass (right) | 0.6375*** | 0.6033*** | **0.6485\*\*\*** | 0.3543*** | 0.2256*** | 0.0894* | 0.0524 |
| Arm fat mass (right) | 0.6319*** | 0.5907*** | **0.6468\*\*\*** | 0.3482*** | 0.2320*** | 0.0946* | 0.0620 |
| Trunk fat mass | **0.5884\*\*\*** | 0.5828*** | 0.5813*** | 0.3224*** | 0.2241*** | 0.0909* | 0.0597 |
| Mother’s age at death | **-0.8209\*\*\*** | -0.7600*** | -0.8071*** | -0.6743*** | -0.3443*** | -0.2122* | -0.0848 |
| Father’s age at death | -0.7392*** | -6648*** | **-0.7438\*\*\*** | -0.6395*** | -0.3414*** | -0.2632*** | -0.1572** |
| Current tobacco smoking | 0.7153*** | **0.7629\*\*\*** | 0.6069*** | 0.6220*** | 0.2266*** | 0.0285 | 0.0024 |
| Cancer | 0.2686*** | **0.3868\*\*\*** | 0.1333* | 0.1300 | 0.1688 | 0.1812 | 0.0228 |
| Heart attack | 0.5115*** | 0.4129*** | **0.5925\*\*\*** | 0.3010** | 0.2735** | -0.0139 | -0.0468 |
| Heart failure | 0.4926* | 0.4288*** | **0.5669\*** | - | - | - | - |
| Stroke | 0.8127*** | 0.6951*** | **0.8483\*\*\*** | - | - | - | - |
| Angina | 0.5694*** | 0.4372*** | **0.6609\*\*\*** | 0.2488** | 0.1706* | 0.0331 | -0.0390 |
| Hypertension | 0.4666*** | 0.3507*** | **0.6037\*\*\*** | 0.1855*** | 0.2328*** | 0.0673 | 0.1209** |

## 4 Discussion

In this paper, we develop and validate a new approach, ENABL Age framework, to measure and interpret biological age using a complex ML model and XAI methods. Our results show that our ENABL Age clock accurately reflects an individual’s health status, predict mortality, and capture various aging mechanisms. To make the ENABL Age clocks explainable, we extend the XAI method, TreeExplainer, to calculate the contributions of input features to ENABL Age in units of year. To improve the applicability of ENABL Age clocks, we train ENABL Age-L for medical professionals using popular blood tests and ENABL AgeQ for non-professional healthcare consumers using questionnaire-based features. Association analysis with risk factors and aging-related morbidities, GWAS results on the ENABL Age clocks trained on different mortality causes show that the ENABL Age clocks with different age-related outcomes capture different aging mechanisms. Our ENABL Age clocks are available on an interactive website (https://suinleelab.github.io/ENABLAge) which allows users to calculate and interpret their own ENABL Age.

Interpretability is extremely important for confidence and utility of biological age clock. Understanding the contributions of features to biological age can enhance scientific knowledge of the aging process, help unhealthy individuals adjust their lifestyles, and accelerate the development of drugs that target aging-related diseases. Complex ML models (e.g., tree-based models, neural networks) can capture high-dimensional and non-linear relationships among features and have shown great predictive performance in a variety of applications. However, they are more difficult to interpret, so most of the previous biological age clocks were based on linear models.

Our ENABL Age approach overcomes this difficulty by adopting XAI techniques. We explain ENABL Age by decomposing ENABL AgeAccel to the contributions of the input features (i.e., feature attributions) in units of years. Our explanations are individualized and can capture various aging processes for different individuals, helping users identify their most important risk factors and adjust their lifestyles. In addition, rescaling the risk scores and feature contributions to the scale of age, especially in units of year, makes them easier to understand because the scale of risk scores may not be meaningful without comparing them to other individuals’ scores; further, a model’s prediction may not be meaningful for non-ML practitioners, but an age acceleration is naturally interpretable for doctors and consumers alike.

ENABL Age is a flexible approach in terms of input features, the age-related phenotype or outcome to predict, and the model. For input features, we use complex ML models to model high-dimensional data with non-linear relationships without the assumptions that linear models require. Furthermore, unlike other second-generation biological age clocks, which carefully select the features related to age-related outcomes prior to model training [25, 26, 31, 51], we incorporate the age-related outcome by directly predicting it, which helps us avoid the complex feature selection step. Using all available features helps us capture more complex and comprehensive aging signals. Also, without the constraint of feature selection, we can easily create models that depend on specific types of features (questionnaire, lab tests, etc.).

Many previous biological age clocks have been built by analysis of omics data, such as epigenetic data [16, 18, 2, 31], transcriptomic data [39, 11, 36], proteomic data [35, 47, 24], etc. Due to its flexibility, we can easily extend ENABL Age to omics data by using these data types as the input features. Moreover, ENABL

Age lets us determine what aging signals to capture by defining different age-related prediction tasks, which can be any age-related phenotypes or outcomes. We ensure the applicability of the ENABL Age approach to other age-related outcomes by predicting dementia using the ROSMAP dataset and validating that ENABL Age accurately captures dementia-related aging signals (Supplementary Appendix Section 1). Hence, we can measure different biological ages for different aspects of aging, achieving a comprehensive understanding of aging processes and answering targeted questions (e.g., what features are important to circulatory system-related aging?).

In terms of the model, we can use other complex ML models instead of GBTs. In this paper, we adopt GBTs because they achieve the best performance on tabular datasets, especially mortality predictions, as shown in previous work [33, 40]. Deep neural networks perform very well when applied to large-scale data in biomedical fields [9, 7, 44] and have been used to measure biological ages using various omics data [14, 43, 13]. However, these models are less informative and useful due to their lack of interpretability. The rapid development of feature attribution methods for neural networks can help overcome this problem [46, 20, 6].

In our future work, the flexibility of ENABL Age will enable us to adopt neural networks and related feature attribution methods to measure and interpret ENABL Age clocks using multi-omic data and multitask learning, which is not feasible using GBTs. Multi-omic data can help identify which aging biomarkers are shared between omics layers or carry distinct information, and multi-task learning allows us to simultaneously measure biological age for different aspects of aging.

A limitation of our study is that ENABL Age does not provide causal insights. This is not unique to our method and poses an essential problem in previous biological age models [42]. However, ENABL Age explanations can surface meaningful biological associations. Moreover, a recent study shows that the first-generation biological age models perform worse than random chance at identifying causal biomarkers; however, methods that incorporate additional information on mortality, such as PhenoAge, can identify potential causal factors of aging with much greater frequency [37]. Since ENABL Age clocks incorporate additional aging phenotypes and outcomes, they similarly has a greater chance of identifying potential causal aging factors. Another study limitation is that we do not apply our approach on omics data, especially DNA methylation data, the most popular type of data used in previous studies to derive biological ages. However, our flexible ENABL Age approach can be easily extended to various types of omics data in future work.

In summary, we develop and validate a new approach, ENABL Age, to measure and interpret biological age related to different aging mechanisms using the combination of a complex ML model and XAI methods. ENABL Age takes a consequential step towards applying XAI to interpret biological age models. Its flexibility allows for many future extensions to omics data, even multi-omic data, and multi-task learning.

## Data Availability

The study used openly available human data from NHANES (https://www.cdc.gov/nchs/nhanes/index.htm) and UK Biobank (https://www.ukbiobank.ac.uk/)

## Footnotes

1 https://www.ukbiobank.ac.uk/

2 http://www.cdc.gov/nchs/nhanes.htm

