## Supplementary Appendix for "An explainable AI framework for interpretable biological age"

### 1 ENABL Age can be applied to other age-related tasks.

To validate that ENABL Age can be extended to age-related tasks other than mortality, we apply ENABL Age on dementia prediction using the ROSMAP dataset from two longitudinal aging cohort studies. We generate samples with no dementia history and have a sample size of 9,103 samples aged 56 to 106 derived from 1,597 individuals. We train a GBT classification model to predict imminent dementia onset (i.e., a diagnosis within the next three years) while learning the ENABL Age and then fit exponential curves between chronological ages and dementia predictions (Supplementary Figure 1A).

In Supplementary Figure 1B, we show that the ENABL Ages of the dementia-onset samples are generally higher than their corresponding chronological ages and the ENABL Ages of the normal samples. Supplementary Figure 1C shows the Kaplan-Meier curve of dementia onset for the individuals, stratified into the healthy (lowest 25%) and unhealthy (highest 25%) agers according to the ENABL Age accelerations of the first visits. The unhealthy agers have much steeper declines in dementia-onset over the 16 years of follow-up, suggesting the dementia ENABL Age's effectiveness on identifying high dementia risk individuals. These results demonstrate that the dementia ENABL Age reflects individuals' dementia risk comparing with the samples of the same age and suggests that the dementia ENABL Age may capture the aging signal related to dementia.

We also perform GWAS on dementia ENABL Age to validate that the dementia ENABL Age captures demential status. Supplementary Figure 1D shows the Manhattan plot and the selected mapped genes of the lead SNPs. All three mapped genes, APOE, APOC1 and NECTIN2, are associated with Alzheimer's disease, indicating that the dementia ENABL Age captures the aging signal related to dementia/Alzheimer's disease, further indicating that ENABL Age can be successfully applied to age-related task beyond mortality.

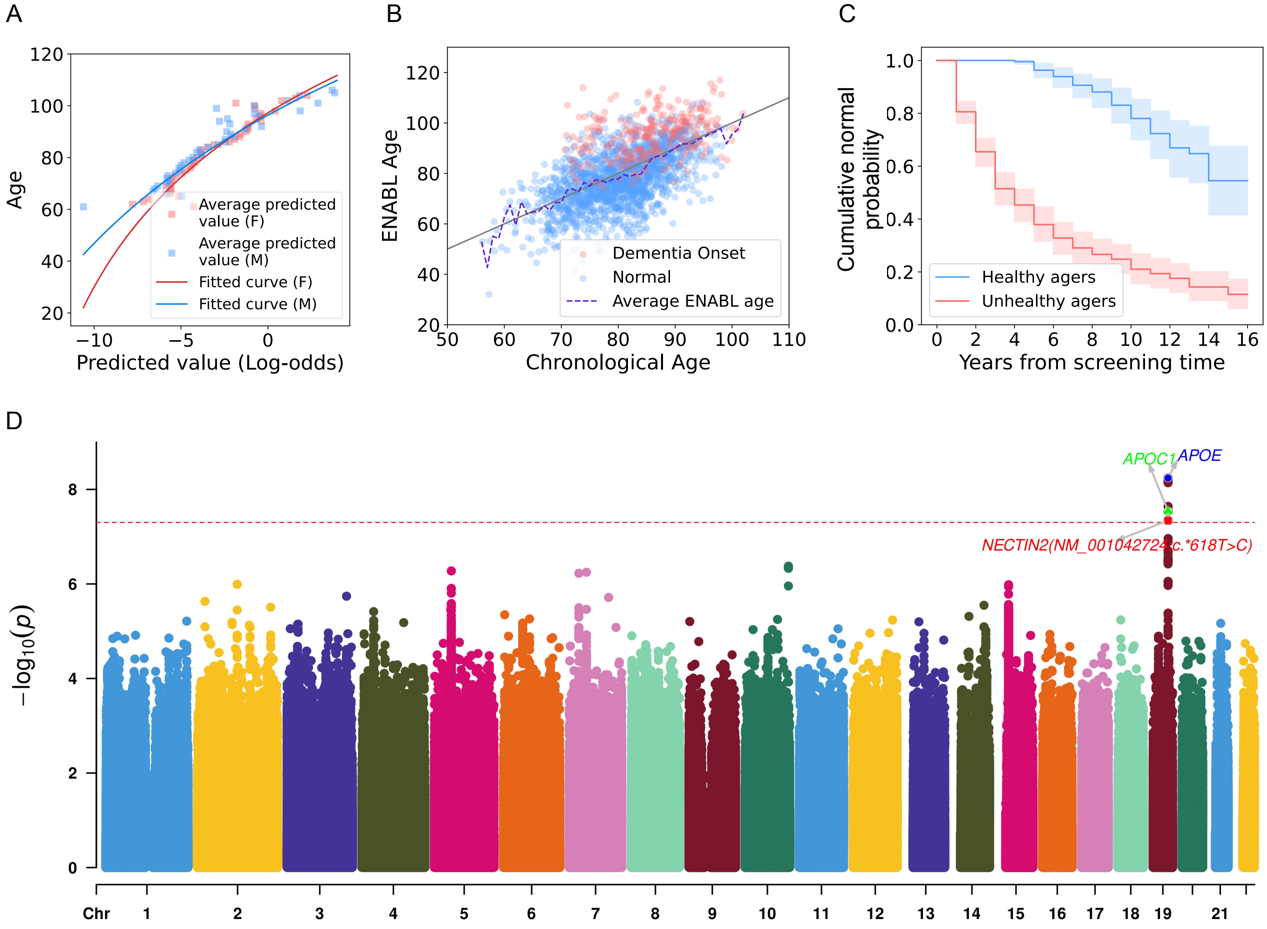


Supplementary Figure 1: **The results of ENABL Age framework on the ROSMAP dataset.** (A) The curves that transform GBTs' predicted value to ENABL Age predictions on the ROSMAP dataset. (B) The scatter plot of ENABL Age predictions versus chronological ages on the ROSMAP dataset. (C) Kaplan–Meier curves for persons in the highest 25% (unhealthy agers) versus the lowest 25% (healthy agers) of dementia onset ENABL age acceleration in the ROSMAP test set. (D) Manhattan plots for dementia onset ENABL Age.

### UK Biobank

A

| Mortality cause | Testing Set | | Geographical validation set | |
|---|---|---|---|---|
| | Linear models | Gradient Boosted Trees | Linear Models | Gradient Boosted Trees |
| All-cause | 0.7633 | **0.8043** | 0.7828 | **0.8161** |
| Neoplasm-cause | 0.7450 | **0.7764** | 0.7625 | **0.7915** |
| Circulatory Disease-cause | 0.8260 | **0.8472** | 0.8481 | **0.8795** |
| Respiratory Disease-cause | 0.8967 | **0.9230** | 0.8969 | **0.9373** |
| Digestive Disease-cause | 0.8860 | **0.8892** | 0.8710 | **0.8991** |
| Other causes | 0.8006 | **0.8519** | 0.8149 | **0.8540** |

B

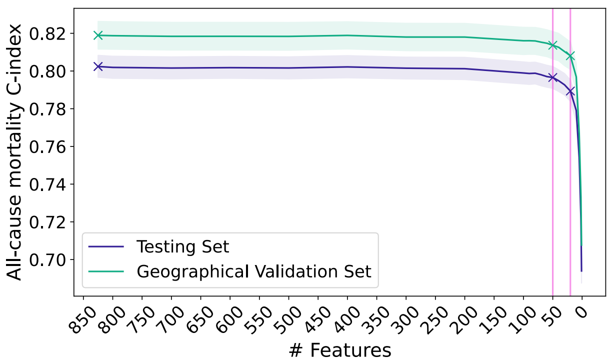


### NHANES

C

| Mortality cause | Linear models | Gradient Boosted Trees | Mortality cause | Linear models | Gradient Boosted Trees |
|---|---|---|---|---|---|
| All-cause | 0.8763 | **0.8834** | Alzheimer's Disease-cause | **0.9723** | 0.9492 |
| Diseases of Heart-cause | 0.9316 | **0.9433** | Diabetes Mellitus-cause | 0.9658 | **0.9698** |
| Malignant Neoplasm-cause | 0.8705 | **0.8786** | Influenza and Pneumonia-cause | **0.9132** | 0.8946 |
| Chronic Lower Respiratory Disease-cause | 0.9694 | **0.9714** | Nephritis, Nephrotic Syndrome and Nephrosis-cause | 0.9544 | **0.9667** |
| Cerebrovascular Disease-cause | **0.8965** | 0.8929 | Other causes | 0.8761 | **0.8912** |

D

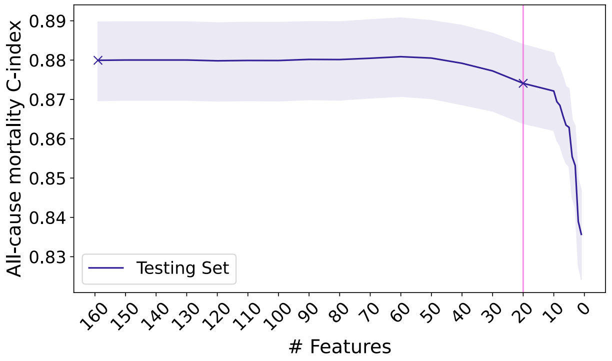


Supplementary Figure 2: (A,C) The C-index of the all-cause mortality prediction and cause-specific mortality prediction on UK Biobank and NHANES datasets using linear Cox regression models, GBTs Cox regression models. (B,D) The C-index of the models using different feature sets after recursive feature elimination on UK Biobank and NHANES datasets.

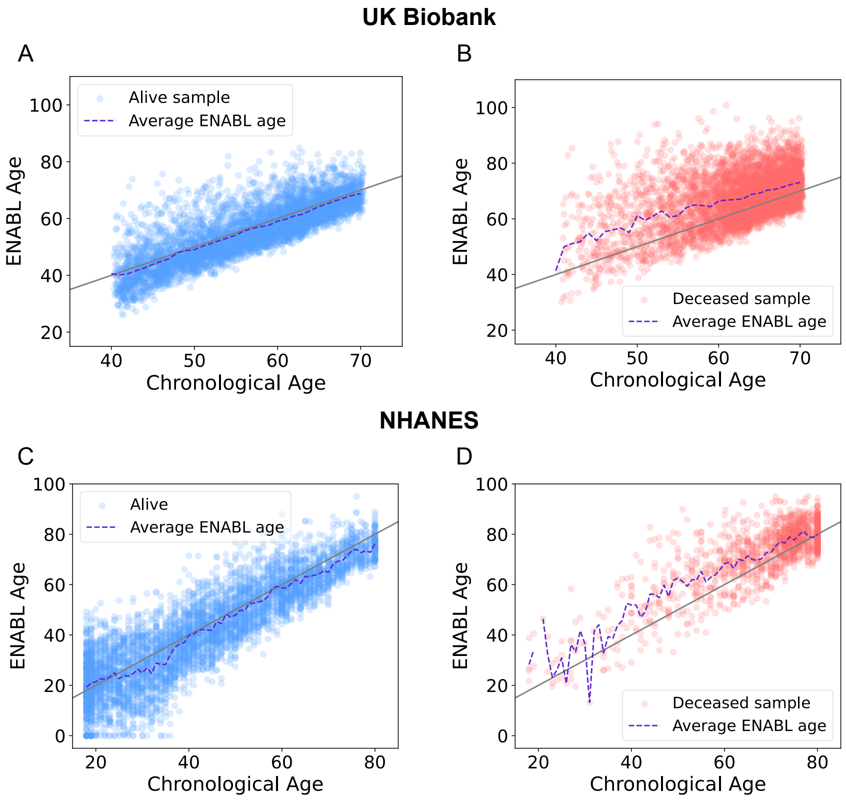


Supplementary Figure 3: (A-D) The scatter plots of ENABL Age predictions versus chronological ages for the individuals that are alive or deceased at mortality data collection time in the UK Biobank and NHANES dataset.

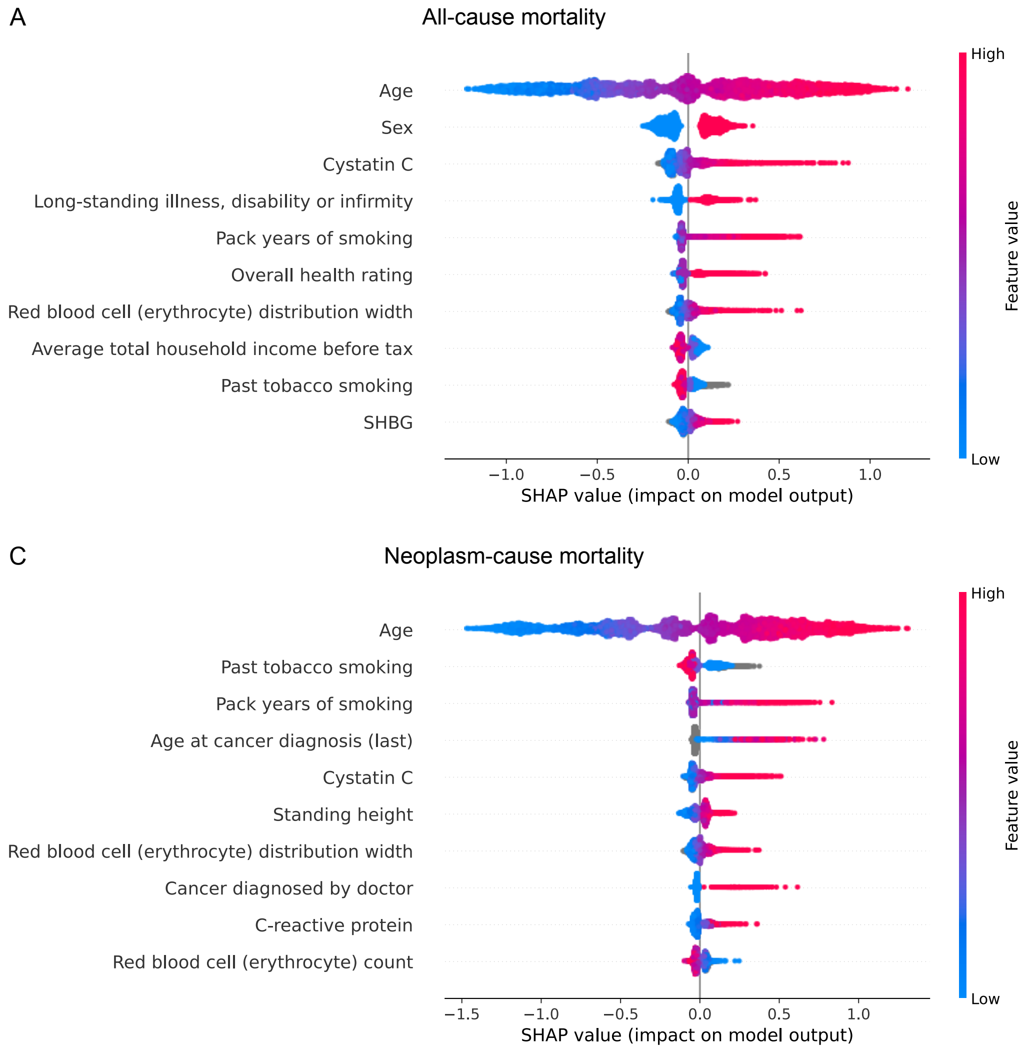


Supplementary Figure 4: (A-B) SHAP summary plot for all-cause mortality and neoplasm-cause mortality prediction models using UKB datasets.

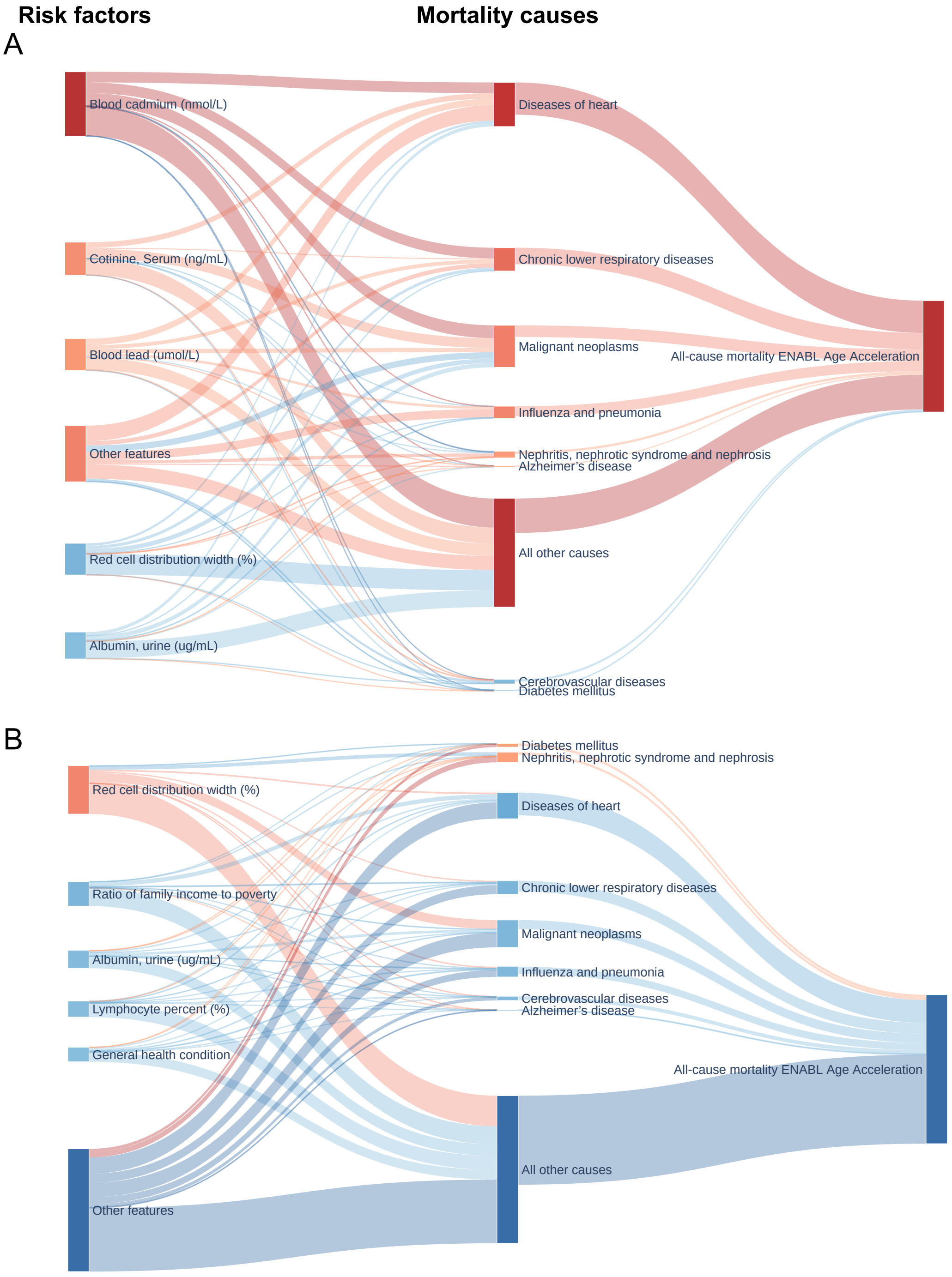


Supplementary Figure 5: (A-B) The two-layer all-cause mortality ENABL Age explanations for two samples from NHANES dataset. The lines in red indicate positive rescaled SHAP values that increase the ENABL AgeAccel; those in blue indicate negative rescaled SHAP values that decrease the ENABL AgeAccel.
