## Supplementary Methods for "An explainable AI framework for interpretable biological age"

### Supplementary Method

#### Methods 1 Data collection and processing

##### Methods 1.1 UK Biobank

The participants were enrolled in the UK Biobank from April, 2007, to July, 2010, from 21 assessment centres across England, Wales, and Scotland using standardised procedures. Ethics approval for the UK Biobank study was obtained from the North West - Haydock Research Ethics Committee (21/NW/0157). Informed consent was obtained from all UK Biobank participants (the consent form is available at `https://www.ukbiobank.ac.uk/consent`). The participants visited their closest assessment centre to provide baseline information, physical measures, and biological samples. In this study, we include all measurements available on November 19, 2020. We exclude (1) features that are missing in more than 80% of the samples and (2) one feature from pairs of highly correlated features with correlations greater than 0.98. After preprocessing, our UKB data has 501,366 samples aged 40-70 with 825 features from numerous categories: demographics, blood assays, health and medical history, lifestyle and environment, physical measures, etc. We use the 35,735 samples from two Scottish centers (Edinburgh and Glasgow) as the geographical validation set. We impute missing data using MissForest [13], a nonparametric random forest-based multiple imputation method for mixed-type data.

The mortality data we use includes all deaths occurring before March 8, 2022. Detailed information about the mortality data is available at `https://biobank.ctsu.ox.ac.uk/crystal/crystal/docs/DeathLinkage.pdf`. As with Ganna and Ingelsson [7], we defined six cause-specific mortality categories using the International Classification of Diseases, edition 10 (ICD-10), classification as follows: neoplasms, C00–D48; diseases of the circulatory system, I05–I89; diseases of the respiratory system, J09–J99; diseases of the digestive system, K20–K93; external causes of mortality and morbidity, V01–Y84; and other diseases, all remaining ICD-10 codes. We only consider the primary cause of death for all participants. The demographic characteristics and sample size of the data for different tasks are shown in Table 1.

##### Methods 1.2 NHANES

The National Health and Nutrition Examination Survey (NHANES) from the National Center for Health Statistics (NCHS)(`http://www.cdc.gov/nchs/nhanes.htm`) conducts interviews and physical examinations to assess the health and nutrition data for all ages in the United States. The interviews include demographic, socioeconomic, dietary, and health-related questions. The examinations include medical, dental, physiological measurements, and laboratory tests administered by highly trained medical personnel. Since 1999, data were collected and released at 2-year intervals. Each year NHANES examines a nationally representative sample of roughly 5,000 individuals across the Unites States. In this study, we include NHANES data collected between 1999 and 2014. We exclude participants under age 18 because they are not eligible for public release mortality data. Our study includes samples with known mortality status who participated in NHANES 1999-2014 ($n = 47,261$). In the raw data, individuals 85 and over are topcoded at 85 years of age in NHANES 1999-2006 and individuals 80 and over are topcoded at 80 years of age in NHANES 2007-2014. To keep consistency, we topcode individuals 80 and over at 80 years of age. We include all demographic, laboratory, examination, and questionnaire features that could be automatically matched across different NHANES cycles. We exclude variables that are missing for more than 50% of the participants and one feature from pairs of highly correlated features with correlations greater than 0.98; after filtering and one-hot encoding, 158 features remain. We also impute missing data using MissForest [13].

All-cause mortality is ascertained by a linked NHANES mortality file that provides follow-up mortality data from the date of survey participation through December 31, 2015. For NHANES 1999-2014, the nine cause-specific death categories in the linked mortality files include the following groups selected from the NCHS underlying cause-of-death recodes: heart disease, cancer (malignant neoplasms), chronic lower respiratory disease, unintentional injuries, cerebrovascular diseases, Alzheimer's disease, diabetes, pneumonia and influenza, and kidney disease. The demographic characteristics and sample size of the data for different tasks are shown in Table 1.

##### Methods 1.3 ROSMAP

The Religious Orders Study (ROS) [1] and Memory Aging Project (MAP) [2] are complementary epidemiological studies that each enroll persons without dementia who agree to annual evaluations and eventual organ donation. ROS enrolls clergy living communally from 40 Catholic groups across the US. As a complementary study, MAP recruited participants from a wider range of life experiences throughout northeastern Illinois. Clinical data collection procedures were consistent between both studies to allow the data to be merged for analyses [2]. Due to their recruitment strategies, followup rates of survivors reached around 95% for both studies.

We use the data from Beebe-Wang et al. [3]. Our prediction task is dementia onset in individuals with no history of dementia within the next three years. We have a sample size of 9,110 samples, of which 1,244 are labeled as positive (dementia onset within the next three years). The data is derived from 1,597 individuals, of which 521 developed dementia. We use 53 features including demographics features, medical history features, lifestyle factors, cognitive tests and genetic features (APOE genotype).

#### Methods 2 ENABL Age approach

We construct ENABL Age clocks in two stages. First, we develop predictors of all-cause mortality and cause-specific mortality on all or a subset of the variables using Cox proportional hazard (CoxPH) models. Second, we fit an exponential curve to the training samples' chronological ages and the predictions of the GBT models. Then, we use the inverse function of the exponential curve to calculate our ENABL Age given a prediction. We define ENABL Age acceleration (AgeAccel) as the difference between the ENABL Age estimate and chronological age.

##### Methods 2.1 Age-related outcome prediction models

To model mortality, we use gradient boosted trees (GBTs) with the Cox regression objective. GBTs are nonparametric methods composed of iteratively trained decision trees. The final ensemble of trees captures non-linearity and interactions between predictors. The dataset is randomly divided into training (64%), validation (16%) and testing (20%) sets. The hyperparameters are chosen by GridSearch using the training and validation set. We use the XGBoost implementation [**chen2016xgboost**] (`https://xgboost.readthedocs.io/en/latest/python/index.html`). The hyperparameters are chosen from the following values:

- Learning rate: 0.01

- Maximum number of trees: 10,000
- Early stopping rounds: 1,000
- Maximum tree depth: {1, 3, 5, 7, 9}
- Subsampling: {0.2, 0.5, 0.8, 1.0}

Parameter values not specified above are left at their default values.

For comparison, we also train linear Cox's proportional hazard models. We use the implementation Lifelines (`https://lifelines.readthedocs.io/en/latest/fitters/regression/CoxPHFitter.html`). The hyperparameters specified below are chosen by GridSearch using the train and validation set. The hyperparameters are chosen from the following values:

- Penalizer: {0, 0.001, 0.01, 0.1, 1, 10, 100}
- L1 ratio: {0, 0.001, 0.01, 0.1, 1, 10, 100}

Parameter values not specified above are left at their default values.

Models' performance is measured with the concordance index (C-index) which is the proportion of concordant pairs divided by the total number of possible evaluation pairs. We bootstrap the test set for 1,000 times and assess the statistical significance of the difference in C-index for pairs of models. Specifically, we resample with replacement from the test set 1,000 times and compare the models' performance on resampled test sets. We report a p-value which is the percentage of times that the linear Cox proportional hazard model is better than or equal to gradient boosted trees, divided by the number of resampled test sets. All models are built in Python 3.7.

To predict dementia onset using ROSMAP data, we use GBTs with a logistic regression objective, but all other training procedures are same an in the mortality prediction models. Models' performance is measured with AUROC. We also bootstrap the test set 1,000 times when evaluating this model.

#### Methods 2.2 Recursive feature elimination

To find less costly but nearly as accurate models, we select features using recursive feature elimination. Recursive feature elimination works by searching for a subset of features by starting with all features in the training dataset and successively removing features until the desired number of features remains. Firstly, we train a model on the full dataset with all features. Then we rank features by importance (according to mean absolute SHAP values) and remove the least important features. Another model is trained on the resulting feature set, and the process iterates until only the desired number of features are left. Starting from all features, we remove about 5 features in each iteration until only one feature is left. The model's performance is shown in Supplementary Figure 2. Specifically, we (bootstrap) resample with replacement from the test set 1,000 times and report the average and the 95% confidence interval of the C-index.

#### Methods 2.3 Rescaling predictions to ENABL Age

The prediction (i.e., log-odds) of the mortality/dementia onset prediction models can be interpreted as a version of ENABL Age which is not contextualized in terms of chronological age. To do so, we non-linearly transform the models' predictions to produce biological age estimates, which are in units of years. Specifically, we fit an exponential curve, with parameters $a$, $b$, and $c$:

$$pred = e^{a \times age + b} + \min(pred) + c,$$

$$c = -0.1, \text{ if } c \geq 0.$$

to the training samples' chronological ages ($age$) and the predictions ($pred$) of the GBT models using the Levenberg-Marquardt algorithm [8]. Here, $\min(pred)$ is the minimum value of the prediction of training samples. We use the inverse function of the exponential curve:

$$\text{ENABL Age} = \frac{\ln(pred - \min(pred) - c) - b}{a}$$

to calculate our ENABL Age given a prediction.

#### Methods 2.4 All-cause mortality ENABL Age prediction using cause-specific ENABL Ages

To obtain the contribution of different cause-specific ENABL Age estimate to the all-cause ENABL Age estimate, we train GBTs models to predict the all-cause mortality ENABL Age estimate using chronological age and cause-specific mortality ENABL Age estimates for different sex using UKB and NHANES dataset. We use GBTs with the mean squared error objective. The dataset is randomly divided into training (64%), validation (16%) and testing (20%) sets. We use the following parameters:

- Learning rate: 0.01
- Maximum number of trees: 1,000
- Early stopping rounds: 100

Parameter values not specified above are left at their default values.

#### Methods 2.5 Interpretation of ENABL Age

To interpret ENABL Age estimate, we utilize TreeExplainer [11], which provides a local explanation of the impact of input features on individual predictions. TreeExplainer calculates exact SHAP [10] (SHapley Additive exPlanations) values for tree-based models.

##### SHAP (SHapley Additive exPlanation) values

Firstly, we calculate the SHAP values for the mortality prediction models. SHAP (SHapley Additive exPlanation) values attribute to each feature the change in the expected model prediction when conditioning on that feature. The change of the model's prediction when the feature is masked is recorded across all possible subsets of features, yielding an average change in prediction resulting from the inclusion of a feature in the model:

$$\phi_i(f, x) = \sum_{R \in \mathcal{R}} \frac{1}{M!} \left[ f_x(P_i^R \cup i) - f_x(P_i^R) \right], \tag{1}$$

where $\phi_i$ is the feature attribution (SHAP value) of feature $i$ in model $f$ for data point $x$, $\mathcal{R}$ is the set of all feature permutations, $P_i^R$ is the set of all features before $i$ in the ordering $R$, $M$ is the number of input features. In this paper, we use baseline shapley values and marginal shapley values. For baseline shapley values ($\phi(f, x^e, x^b)$), $f_x(S) = f(x^e_S, x^b_{\bar{S}})$, where $f(x^e_S, x^b_{\bar{S}})$ denotes evaluating $f$ on a hybrid sample where present features are taken from the explicand $x^e$ and absent features are taken from the baseline $x^b$. Then we estimate marginal Shapley values by first estimating baseline Shapley values for many baselines and then averaging them [6].

SHAP values guarantee a set of desirable theoretical properties, including additivity and consistency. In particular, additivity states that when approximating the original model $f$ for a specific input $x$, the SHAP values sum up to the output $f(x)$:

$$f(x) = \phi_0(f) + \sum_{i=1}^{M} \phi_i(f, x), \tag{2}$$

where $\phi_0(f) = \mathrm{E[f(x)]} = \mathrm{f_x}(\emptyset)$ that is the average model prediction on the baseline samples. Consistency states that if a model changes so that some feature's contribution increases or stays the same regardless of the other inputs, that input's attribution should not decrease.

For the age-related outcome (e.g., mortality and dementia onset) prediction models, we use the baselines and explicands (i.e., samples being explained) of the same age and sex. The baselines (i.e., background samples) are from the training set and the explicands are from the testing set. Therefore, the $\phi_0(f)$ for a specific age equals to the average predictions of the samples of that age in the training set.

##### Rescaling SHAP values to the ENABL Age space

We rescale the SHAP values to the ENABL Age space so that the rescaled SHAP values are in units of years and sum to the ENABL AgeAccel. To do so, we use the generalized rescale rule proposed in our previous work [5]. Suppose we have a age-related outcome prediction model $f$, and a non-linear model $g$ that rescales the prediction to ENABL Age. We can calculate ENABL Age using $h(x) = g(f(x)), x \in \mathbb{R}^m$. We first calculate the baseline shapley values $\phi(f, x^e, x^b)$ for the age-related outcome prediction model $f$ given an explicand $x^e$ and a baseline $x^b$. Then, based on the generalized rescale rule, we can calculate the baseline shapley values for $h$ as follows:

$$\phi(h, x^e, x^b) = \phi(f, x^e, x^b)((h(x^e) - h(x^b)) \oslash (f(x^e) - f(x^b))),$$

where $\oslash$ denotes Hadamard division. After rescaling, the $\phi_0(h)$ for a specific age equals to the average ENABL Age of the samples of that age in the training set, which is approximate to that age. At the end, we average the baseline shapley values to produce an estimate of the SHAP values (marginal shapley values). Therefore, the sum of the rescaled SHAP values is approximate to the ENABL Age acceleration. As a consequence, we can decompose the ENABL Age to the contribution of different features in units of years.

##### Two-layer explanation of all-cause mortlality ENABL AgeAccel

We train models to predict the all-cause ENABL Age estimates using chronological age and cause-specific ENABL Age estimates for different sexes, so that we can rescale the SHAP values of different cause-specific mortality predictions to all-cause mortality ENABL AgeAccel. We use $t$(chronological age, $h_0(x)$, $h_1(x)$, $\cdots$, $h_{k-1}(x)$) to denote the all-cause ENABL Age prediction model, where $k$ is the number of mortality causes we consider. We can rescale the SHAP values as follows:

$$\begin{aligned}\phi(t, x^e, x^b) &= \phi(h, x^e, x^b)(\phi(t, x^e, x^b) \oslash (h(x^e) - h(x^b))) \\ &= \phi(f, x^e, x^b)(\phi(t, x^e, x^b) \oslash (f(x^e) - f(x^b)))\end{aligned}$$

Consequently, we achieve the two-layer explanations of ENABL AgeAccel, i.e., the features to different cause-specific mortality as well as cause-specific mortality to all-cause mortality ENABL Age estimates.

### Methods 3 5-year and 10-year mortality prediction models

#### Methods 3.1 UK Biobank and NHANES mortality prediction models

In Section 3.2, we directly evaluate the mortality prediction power of ENABL AgeAccel, PhenoAgeAccel and BioAgeAccel by training 5- and 10-year mortality prediction models adjusted by chronological age and sex. Firstly, We calculate PhenoAge and BioAge using the formulae in their original papers. Then, we train all-cause mortality ENABL Age clocks using the features included in PhenoAge and BioAge to show the effectiveness of ENABL Age framework. We further build all-cause mortality ENABL Age clocks using other subsets of features: laboratory features in the four most popular blood panels (ENABL Age-L) – CBC (Complete Blood Count), CMP (Comprehensive Metabolic Panel), LP(Lipoprotein (a)) and WBC (White Blood Cell Count ), the top 20 most important questionnaire features (ENABL Age-Q), and the top 20 most important features (ENABL Age-20). We train GBTs to predict 5- and 10-year all-cause mortality using ENBAL AgeAccel, PhenoAgeAccel, and BioAgeAccel, chronological age and sex. To compare with chronological age, we also train 5- and 10-year mortality prediction models using only chronological age and sex. The dataset is randomly divided into training (80%) and testing (20%) sets. We use GBTs using the logistic regression loss with default hyperparameters. Models' performance is measured with AUROC. We bootstrap the test sets/geographical validation set for 1,000 times and obtain the confidence intervals of the AUROCs.

#### Methods 3.2 Cross dataset validation

We perform a cross-dataset validation using ENABL Age-L by training it on UKB samples using the features in the four popular blood panels that overlap between UKB and NHANES datasets and evaluating the 5- and 10-year mortality prediction power on NHANES testing samples aged 40-70. Specifically, the ENABL Age-L is trained on UKB samples and used to calculate the ENABL Age-L for NHANES samples. Then, we train and test 5- and 10-year mortality prediction models using the ENABL Age-L, chronological age and sex on NHANES samples aged 40-70. For "Train on NHANES, test on NHANES", we train the ENABL Age-L on NHANES training samples aged 40-70 and used it to calculate the ENABL Age-L for NHANES testing samples. Then, we train and test 5- and 10-year mortality prediction models using the ENABL Age-L, chronological age and sex on NHANES samples aged 40-70. For "Train on UKB, test on UKB", we train the ENABL Age-L on UKB training samples aged 40-70 and used it to calculate the ENABL Age-L for UKB testing samples. Then, we train and test 5- and 10-year mortality prediction models using the ENABL Age-L, chronological age and sex on UKB samples. To compare with chronological age, we also train 5- and 10-year mortality prediction models using only chronological age and sex on NHANES samples aged 40-70.

### Methods 4 Association analysis

We examine the associations of ENABL AgeAccels, PhenoAgeAccel, BioAgeAccel with a range of risk factors and age-related morbidity outcomes using UKB test set. We consider the following risk factors and age-related morbidity outcomes:

- Pack years of smoking: Pack years calculated for individuals who have smoked. The general definition of a pack year is the number of cigarettes smoked per day, divided by twenty, multiplied by the number of years of smoking. The number of years of smoking is calculated by substracting the age of starting smoking from the age smoking was stopped. The pack years of smoking is available in data field 20161.
- Walking pace: The walking pace is collected from the touchscreen question ”How would you describe your usual walking pace?” from all participants except those who indicated they were unable to walk. We consider ”less than 4 miles per hour” as slow pace and ”more than 4 miles per hour” as fast pace. The walking pace is available in data field 924.
- Grip strength: We use the maximum for the left hand grip strength and right strength grip strength. The units of grip strength is Kg. The grip length measurements are available in data field 46 (left hand) and 47 (right hand).
- Forced expiratory volume in 1-second: We use the highest measure from the array of values for Forced Expiratory Volume in 1-second which is in units of liter. The forced expiratory volume in 1-second is available in data field 20150.
- Waist-hip ratio: We calculate the waist-hip ratio using waist circumference divided by the hip circumference. The waist circumference is available in data field 48 and the hip circumference is available in data field 49.
- Cancer diagnosis: The time-to-event is calculated by substracting the date of attending assessment centre from date of cancer diagnosis. The date of cancer diagnosis is available in data field 40005.
- Myodardial infarction diagnosis: The date of myodardial infarction diagnosis is available in data field 42000.
- Stroke diagnosis: The date of stroke diagnosis is available in data field 42006.
- COPD diagnsois: The date of COPD diagnosis is available in data field 42016.
- Asthma diagnosis: The date of asthma diagnosis is available in data field 42014.
- All-cause dementia diagnosis: The date of all-cause dementia diagnosis is available in data field 42018.
- End-stage renal disease diagnosis: The date of end-stage renal disease diagnosis is available in data field 42026.

The risk factors and age-related morbidity outcomes measures are regressed separately on each of the age accelerations adjusting for chronological age (years), sex using ordinary least squares regression (pack years of smoking, grip strength, forced expiratory volume in 1-second and waist-hip ratio), logistic regression (walking pace), and Cox regression (cancer diagnosis, myodardial infarction diagnosis, stroke diagnosis, COPD diagnsois, asthma diagnosis, all-cause dementia diagnosis and end-stage renal disease diagnosis) as appropriate. We report the change in each of the outcome measures associated with a year increase in age acceleration, that is beta for ordinary least squares regression, odds ratio for logistic regression and hazard ratio for Cox regression. We use the implementation Statsmodels (`https://www.statsmodels.org/stable/index.html`) for ordinary least squares regression and logistic regression and Lifelines (`https://lifelines.readthedocs.io/en/latest/fitters/regression/CoxPHFitter.html`) for Cox regression. All analyses are conducted using Pythob 3.7.

### Methods 5 Genome-wide association analysis

#### Methods 5.1 Quality control

In our GWAS analysis, we include all white British that have very similar genetic ancestry based on a principal components analysis of the genotypes in UK Biobank, identified using the data field 22006. Additionally, one in third-degree or closer pairs are removed, identified via pairwise kinship coefficients. SNPs are excluded if meeting any of the criteria: (1) missing rates exceeding 0.01 (2) MaCH Rsq imputation quality metric $<$ 0.3, (3) minor allele frequency $< 0.1\%$, (4) Hardy–Weinberg equilibrium test p-value $< 10^{-6}$, (5) missing imputation information score, minor allele frequency, or Hardy–Weinberg equilibrium test result. Overall, 12,755,286 SNPs passed the quality control. The quality control is implemented using PLINK 2.0 `https://www.cog-genomics.org/plink/2.0/`.

#### Methods 5.2 GWAS analysis

The association between ENABL Age accelerations with each SNP is examined using an efficient Bayesian linear mixed effects model (BOLT-LMM software version 2.4; `https://alkesgroup.broadinstitute.org/BOLT-LMM/BOLT-LMM_manual.html`) [9] adjusted for chronological age, sex, genotyping array type, and assessment center, and top 20 genetic principal components. By default, the LD scores included in the BOLT-LMM for European-ancestry samples are used to calibrate the BOLT-LMM statistic. SNP p-values smaller than $5 \times 10^{8}$ are considered to be statistically significant. Manhattan plots are created using the CMplot R package.

We also perform a stepwise model selection procedure on the genome-wide SNP summary statistics to identify independent signals ($p < 5 \times 10^{8}$) using the COJO (Conditional and Joint association analysis) [14] model in the GCTA (Genome-wide Complex Trait Analysis) software (`https://yanglab.westlake.edu.cn/software/gcta/#Overview`) [15]. SNPs more than 10,000 kb away from each other are assumed to be in complete linkage equilibrium. As SNPs are selected, the SNPs with multiple regression R2 greater than 0.9 with already pre-selected SNPs are excluded, so that redundant signals from high LD are excluded. The significant SNPs are mapped to genes based on GRCh37/hg19 coordinates, and were used in searches for published GWAS associations based on GWAS catalog.

#### Methods 5.3 Genetic correlations

We perform cross-trait LD score regression (LDSC) using GWAS summary statistics [4] to relate ENABL AgeAccel to various health-related traits: anthropometric traits (e.g., BMI), adiposity (e.g., whole body/leg/arm/trunk fat mass), longevity (mother/father's longevity), lifestyle (e.g., smoking) and several diseases (e.g., heart attack, heart failure, angina, stoke and hypertension). We use the LDSC (v1.0.1) implementation `https://github.com/bulik/ldsc`. GWAS summary statistics for health-related traits are downloaded from the Ben Neale Lab round 2 `http://www.nealelab.is/uk-biobank`. As recommend by LDSC, we filtered to HapMap3 SNPs for each GWAS summary data, which could help align allele codes of our GWAS results with other GWAS results for the genetic correlation analysis. We also compare with the genetic correlations of GrimAgeAccel, PhenoAgeAccel, Hannum AgeAccel and Horvath AgeAccel which are reported in McCartney et al. [12].
